# Integrating Machine Learning-Based Variable Selection into Heat Vulnerability Index Design

**DOI:** 10.64898/2026.03.29.26349672

**Authors:** Shuyue Qu, Jana Sillmann, Benjamin W. Barrett, Peter M. Graffy, Benjamin Poschlod, Lukas Brunner, Raed Mansour, Malte von Szombathely, Finley Hay-Chapman, Teresa H. Horton, Jennifer Chan, Sheetal Khedkar Rao, Kyra Woods, Abel N Kho, Daniel E. Horton

**Author notes:** Author to whom any correspondence should be addressed., /.

## Abstract

As climate change intensifies, health risks from extreme heat are rising. Accurate assessment of heat vulnerability at high spatial resolution is crucial for developing effective adaptation strategies, particularly in socioeconomically heterogeneous urban settings. However, the identification of key indicators underlying heat vulnerability remains challenging. Using Chicago, Illinois (USA) as a case study, we systematically compare different variable selection strategies in community-level heat vulnerability assessments. We take the conventional unsupervised principal component analysis (PCA)-based Heat Vulnerability Index (HVI) as a baseline, and compare it with supervised approaches that incorporate variable selection, including machine learning algorithms (Lasso regression, Random Forest, and XGBoost) as well as traditional statistical methods (simple linear regression and polynomial regression). Using the vulnerability indicator subsets identified by each variable selection method, we construct multiple HVIs and evaluate their performance against heat-related excess mortality. Our work indicates that supervised variable selection improves the performance of HVIs in capturing heat-related health risks. Among all methods, the Random Forest-based variable selection algorithm achieves the best overall results, highlighting the potential of machine learning to enhance heat vulnerability assessment tools. Our results demonstrate that poverty rate, lack of air conditioning, and proportion of residents aged 65 and above are robust determinants of heat vulnerability in Chicago.

## 1. Introduction

In recent years, climate change has exacerbated the frequency, intensity, and duration of extreme heat events (IPCC, 2023; World Health Organization, 2024; World Meteorological Organization, 2023). Substantial epidemiological evidence suggests that heat is closely linked to increased risks of multiple adverse health outcomes, including cardiovascular and respiratory diseases, renal disorders, adverse pregnancy outcomes, and mental health conditions such as anxiety and depression (Bell et al., 2024; Benmarhnia et al., 2015). Heat-related mortality has been identified as one of the most serious climate threats to human health (Lüthi et al., 2023), and with continued global warming, heat-related morbidity and mortality are projected to rise (Ebi et al., 2021). Given current and impending climate and health challenges, accurate assessments of heat-related health risks are critical for guiding targeted and effective public health interventions and adaptations.

However, not all populations are at equal health risk from heat (Reid et al., 2009). Previous studies indicate that heat vulnerability is associated with a wide range of factors, including socioeconomic conditions, demographic characteristics, and the built environment (Niu et al., 2021; Chuang & Gober, 2015; Reid et al., 2009). Because direct measures of heat vulnerability are unavailable, researchers have developed proxy-based tools, such as Heat Vulnerability Indices (HVIs), that integrate multidimensional indicators of heat-related health risk to identify the populations and communities most vulnerable to heat. HVIs are increasingly used to guide where cities prioritize investments such as cooling access, outreach, housing improvements, and greening initiatives.

One of the most influential HVI frameworks was proposed by Reid et al. (2009), who developed a Principal Component Analysis (PCA)-based HVI at the census tract level for the United States (U.S.). This unsupervised approach applies PCA to pre-selected heat vulnerability indicators to identify latent factors that explain the largest variance without reference to health outcome data. These factors are then interpreted as representing underlying dimensions in heat vulnerability and aggregated to obtain cumulative HVI scores (Cheng et al., 2021; Tate, 2012; Reid, et al., 2009). The Reid et al. methodology has since been widely adopted in heat vulnerability assessments, both in the U.S. and in other global settings (Wang et al., 2023; Manware et al., 2022; Conlon et al., 2020). However, this methodology has certain limitations, including reliance on pre-selected and subjective indicators and lack of validation against health outcomes (Tate, 2012; Cheng et al., 2021; Wolf et al., 2014; Niu et al., 2021), thereby raising concerns about whether unsupervised HVIs adequately capture heat-related health risks.

Building on the unsupervised HVI framework proposed by Reid et al., Conlon et al. (2020) introduced a supervised HVI that incorporates health outcome data in the index construction. This approach linearly regresses each potential vulnerability indicator against the proportion of mortality on extreme heat days and applies the PCA to only those indicators that were moderately significantly associated (p < 0.20) with mortality. However, both the supervised and unsupervised HVIs explained only a small proportion of the variation in mortality on extreme heat days in Detroit, Michigan (Conlon et al., 2020). This limited performance may indicate that variable selection based on simple linear regression cannot fully capture the relationships between heat vulnerability indicators and heat-related health outcomes, suggesting the need to explore nonlinear associations and other modeling approaches. Indeed, a recent study demonstrated that nonlinear models more accurately characterize the relationships between socioeconomic variables and health outcomes compared to linear models (Jeong et al., 2024), which motivates the use of more advanced and flexible variable selection strategies.

In this context, machine learning offers a promising methodological direction for public health research and risk assessment, supporting more targeted interventions (Potash et al., 2015; Sadilek et al., 2018; Potash et al., 2020). While not inherently superior to traditional statistical methods, these approaches can provide locally validated, outcome-informed screening of indicators. Beyond the limitations of simple linear regression, machine learning-based variable selection methods are capable of identifying complex relationships among variables, evaluating variable importance relative to health outcomes during model training, and balancing computational efficiency with overfitting resistance (Guo & Zhou, 2022; Zhao et al., 2023; Zhou et al., 2024; Theng & Bhoyar, 2024). These capabilities suggest that machine learning-based variable selection methods may help refine the indicators used in HVI construction.

This study incorporates machine learning-based variable selection methods into the conventional PCA-based HVI framework to explore the effectiveness of different variable selection strategies. Within the PCA-based HVI framework, three approaches are evaluated and compared: (a) unsupervised HVI using pre-selected indicators from Reid et al. (2009) for PCA; (b) supervised HVI that uses traditional statistical methods for variable selection, including simple linear regression (SLR) (following Conlon et al., 2020) and polynomial regression (PR); and (c) supervised HVI that uses machine learning-based methods for variable selection, including Least Absolute Shrinkage and Selection Operator (Lasso) regression, Random Forest (RF), and XGBoost. We apply these approaches to the 77 community areas (CAs) in Chicago, Illinois, which serves as an ideal study area due to the availability of high-resolution environmental, socioeconomic, demographic, and health data at the CA level. The performance of the HVIs derived from these approaches is systematically validated against CA-level heat-related excess mortality, providing methodological insights and recommendations for future heat vulnerability research. Given that HVI maps influence the prioritization of community resources, this study emphasizes locally justifiable indicators and outcome-informed evaluation, both of which have important equity implications.

## 2. Data

The data for this study are collected and processed at the Chicago CA level using the CA boundaries obtained from the Chicago Data Portal (shown in Supplemental Figure S1). CAs are stable geographical divisions that have been largely unchanged since the 1920s. Many demographic and socioeconomic indicators in Chicago are released and analyzed at this level, providing comparable data across the 77 different CAs over time. Based on these CA boundaries, we integrate mortality and socioeconomic datasets, and aggregate meteorological and land use data to the CA level for subsequent analyses.

### 2.1 Meteorological Data

Daymet Version 4 R1 contains North American daily meteorological data from 1980 to present at a resolution of 1 km. This dataset provides gridded weather estimates using statistical modeling techniques to interpolate and extrapolate ground-based observations (Thornton et al., 2022). It is a research product of the Environmental Sciences Division at Oak Ridge National Laboratory, and widely used in ecosystem modeling and climate change research (Walton & Hall, 2018). We extracted daily minimum temperature (*T_min_*), maximum temperature (*T_max_*), and vapor pressure (V_p_) data for the summer months (May to September) from 1993 to 2019. These daily gridded estimates were then spatially aggregated to the CA level by unweighted averaging of cells whose centroids fall within each CA polygon in Chicago.

### 2.2 Mortality Data

Mortality data used in this study contain individual-level records on age, sex, race, and CA of residence for decedents in Chicago from 1993 to 2019. The dataset also includes the underlying cause of death, coded according to the International Classification of Diseases, Ninth and Tenth Revisions (ICD-9 and ICD-10). These data were provided by the Illinois and Chicago Departments of Public Health (IDPH and CDPH). The use of mortality data was governed by Data Use Agreements with IDPH and CDPH.

### 2.3 Population Data

Population data used in this study were obtained from the standardized time series tables provided by the National Historical Geographic Information System, a project of the Integrated Public Use Microdata Series at the University of Minnesota (Manson et al., 2024). These datasets geo-standardize the 1990, 2000, and 2010 decennial census data to the 2010 census tract boundaries, providing fine-grained population information over time. Annual population estimates were generated using linear interpolation, assuming linear population change between adjacent decennial census years. For the years after 2010 (2011-2019), we used the American Community Survey (ACS) five-year estimates as supplements. All population data were aggregated to the Chicago 2010 CA boundaries to ensure geographic consistency. Chicago population characteristics and all-cause mortality counts for the study period are presented in Table 1.

**Table 1.**
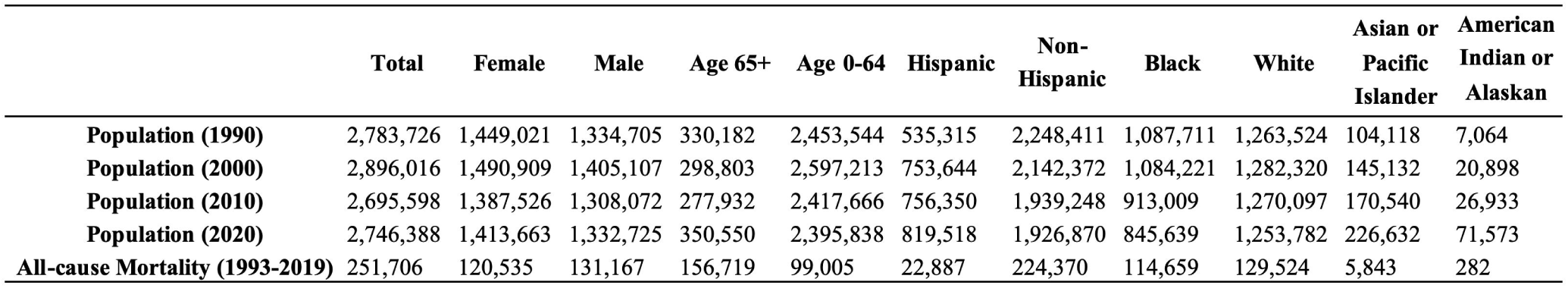
Population characteristics in Chicago for census years 1990, 2000, 2010, and 2020, and total mortality counts for 1993-2019. In the years 2000, 2010, and 2020, some individuals were categorized as belonging to multiple racial groups. To accurately represent the distribution across racial groups, these individuals were counted in each of their respective racial categories; therefore, the sum across racial groups exceeds the total population.

### 2.4 Heat Vulnerability Indicators

Heat vulnerability indicators refer to measurable socioeconomic, demographic, and environmental variables that represent specific dimensions of community-level heat vulnerability and serve as the candidate inputs for constructing the composite Heat Vulnerability Index (HVI). We adapted the ten heat vulnerability indicators proposed by Reid et al. (2009). These indicators were organized into four categories: diabetes prevalence (*Diabetes*), demographic characteristics (*Race other than White*, *Age > 65 years*, *Live alone*, *Age > 65 living alone*, *Below poverty line*, *Less than high school diploma*), land cover (*Not Green Space*), and air conditioning (AC) access (*No central AC*, *No AC of any kind*). Due to differences in data availability and measurement across datasets, the operationalization of several indicators differs slightly between the original Reid et al. (2009) framework and our work (detailed crosswalk is provided in Supplementary Table S1).

Specifically, the diabetes indicator was represented by *Diabetes Prevalence*, obtained from the annual Healthy Chicago Survey conducted by the CDPH. This indicator is defined as percent of adults who reported that they have ever been diagnosed with diabetes by a doctor, nurse or other health professional, excluding pre-diabetes and diabetes only during pregnancy.

Data on age, race, and educational attainment were obtained from the Community Data Snapshots provided by the Chicago Metropolitan Agency for Planning. To better reflect racial and ethnic disparities in heat vulnerability, we split the “*Race other than White*” indicator into two specific indicators: the proportion of Non-Hispanic Black residents (*Black (Non-Hispanic)*) and the proportion of Hispanic or Latino residents (*Hispanic or Latino (of Any Race)*). Proportions of the population aged 65 and over (*Age above 65*), individuals living alone (*Living Alone*), older adults living alone (*Age above 65 and Living Alone*), and educational level (*Less than High School Diploma*) were defined in alignment with Reid et al. (2009). *Poverty Rate* was derived from the CDPH Chicago Health Atlas platform and is defined as the proportion of residents living in households below the federal poverty threshold. These indicators were based on the ACS five-year census tract level estimates and are reported as percentages at the CA level.

For the vegetation indicator *Not Green Space*, we used land cover data from the National Land Cover Database (NLCD) covering the Chicago area for the years 1993-2022. The NLCD is a land cover dataset derived from Landsat satellite imagery, with a spatial resolution of 30 m. We aggregated the land cover data for each year at the CA level. Each pixel was assigned to the CA polygon in which its centroid was located. Following the classification used by Reid et al. (2009), green space is defined as the area of deciduous forest, evergreen forest, mixed forest, dwarf scrub, orchards/vineyards/other, pasture/hay, small grains, fallow, row crops, urban/recreational grasses, palustrine forested wetlands, and palustrine scrub/shrub wetlands. The percentage of green space for each CA was calculated as the total area of these land cover classes divided by the total CA area. The percentage of *Not Green Space* was then calculated as 100 minus the percentage of green space. Finally, a multi-year average of *Not Green Space* was computed for each CA over the period 1993–2022 to represent long-term non-green space coverage. The AC access indicator (*No AC Access*) was obtained from the Healthy Chicago Survey, which for the first time in 2023 included a question on household AC availability: “Is any air conditioning equipment used in your home?”. The questionnaire results provide the proportion of “yes” and “no” responses. To ensure consistent variable directionality, *No AC Access* was calculated as 100 minus the percentage of AC access within each CA. Given the lack of available data on central AC at the CA level, and because the survey question captures all forms of AC including central systems, *No AC Access* was adopted as the only indicator of overall AC access in this study.

In summary, 10 heat vulnerability indicators were included in our analysis (Table 2). All indicators were averaged over their available data periods. Correlations among these heat vulnerability indicators are presented in Supplemental Figure S2.

**Table 2.**
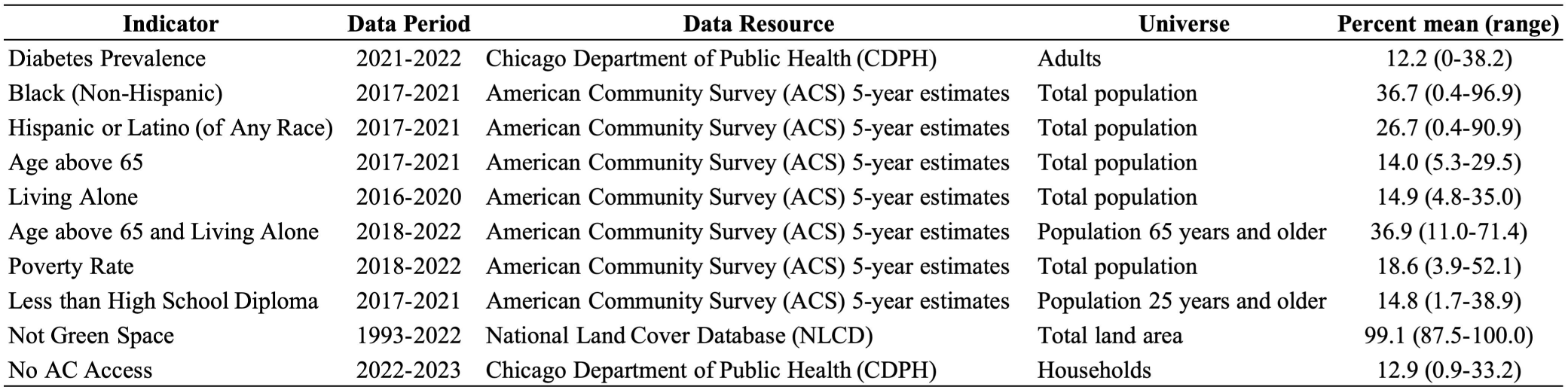
Data period, resources, and ranges for the heat vulnerability indicators used in this study.

## 3. Methods

### 3.1 Identification of Heat Days

Previous studies have shown that the most severe health impacts of heatwaves often happen during nighttime heat exposure (Sarofim et al., 2016). The Fifth National Climate Assessment (U.S. Global Change Research Program, 2023) defines warm nights as days with minimum temperatures at or above 70°F (21.1°C), which provides a reference threshold. We used the daily minimum Heat Index (HI) instead of air temperature as the exposure metric. The HI combines air temperature (T) and relative humidity (RH) and can better characterize the “feels like” temperature of the human body.

The *HI_min_* for each CA was calculated based on the daily minimum air temperature (*T_min_*) and relative humidity (RH) following the NWS Rothfusz regression equation (NOAA, 2022). To determine the RH input, we first computed a weighted daily mean temperature (*T_mean_*) as 0.606×*T_max_* + 0.394×*T_min_* (Spangler et al., 2019). Saturated vapor pressure (*e_s_*, in Pa) was then derived from *T_mean_* using a Magnus-Tetens formula:

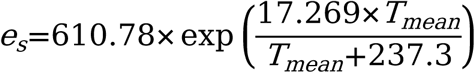

RH (%) was calculated as the ratio of actual vapor pressure (V_p_) to *e_s_*:

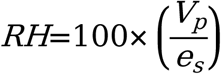

The resulting RH was applied to *T_min_* to compute the raw *HI_min_*. To capture the acute effect of heat, a two-day moving average was applied to these *HI_min_* values (Conlon et al., 2020; Barnett and Åström 2012). A “heat day” was defined as a day when *HI_min_* exceeded 70°F (21.1°C) for at least two consecutive days. This approach captures both the immediate physiological impact of short-term heat exposure and the sustained nature of extreme heat events.

### 3.2 Heat-Related Excess Mortality

We estimated heat-related excess mortality by defining the observed number of deaths (*O*) as the daily average all-cause mortality during the index period (heat days) and the expected number of deaths (*E*) as the daily average mortality during the reference period (non-heat days). Excess mortality was then calculated as *O−E*(Fouillet et al., 2006; Stang et al., 2020; Azhar et al., 2014). Analyses were restricted to the hot season (May to September) for each year. To avoid interference from the COVID-19 pandemic, the time window for heat-related excess mortality estimation was limited to 1993–2019. Annual average heat-related excess mortality was calculated by multiplying the daily values by the total number of heat days in the corresponding year. Annual and daily time series of heat-related excess mortality were generated to examine temporal trends. Heat-related excess mortality was mapped at the CA level, and Moran’s I was calculated to evaluate spatial autocorrelation.

### 3.3 Unsupervised HVI

The PCA-based HVI framework proposed by Reid et al. (2009) is an unsupervised approach that generates HVI scores from a set of pre-selected heat vulnerability indicators. PCA is a dimensionality reduction method that transforms multiple potentially correlated variables into a smaller set of uncorrelated components, capturing most of the variance in the original data (Li et al., 2025; Mallen et al., 2019; Sun et al., 2017). It is a popular approach in the construction of many societal vulnerability indices (Tate, 2012).

In this framework, we perform the PCA on the heat vulnerability indicators using Python’s Scikit-learn PCA tool. Varimax rotation is used to ensure the orthogonality among principal components. Following the approach of Reid et al. (2009), the number of principal components retained is based on a combination of eigenvalues greater than one, significant breaks in the scree plot, and a cumulative explained variance exceeding 75%. We refer to these retained principal components as “factors.” Subsequently, each CA is assigned a score from 0 to 6 for each component, where 0 is assigned to CAs with values ≥ 2 standard deviation (SD) below the mean, and 6 is assigned to CAs with values > 2 SD above the mean (Chuang & Gober, 2015). Factor scores, equally weighted, are summed to calculate the cumulative HVI scores for each CA in Chicago.

### 3.4 Supervised HVI

For supervised approaches, the same candidate indicators from the unsupervised approach were used, and heat-related excess mortality was added as the response variable. Compared to the unsupervised approach, supervised HVI involves a data-driven variable selection step, where heat vulnerability indicators are the predictors and heat-related excess mortality is the health outcome. This study integrates three machine learning-based variable selection methods (Lasso regression, RF, and XGBoost) into the PCA-based HVI framework, and compares them with the results derived from the unsupervised HVI and HVIs that use more traditional supervised variable selection methods, i.e., simple linear regression (SLR) and polynomial regression (PR) (Figure 1).

**Figure 1.**
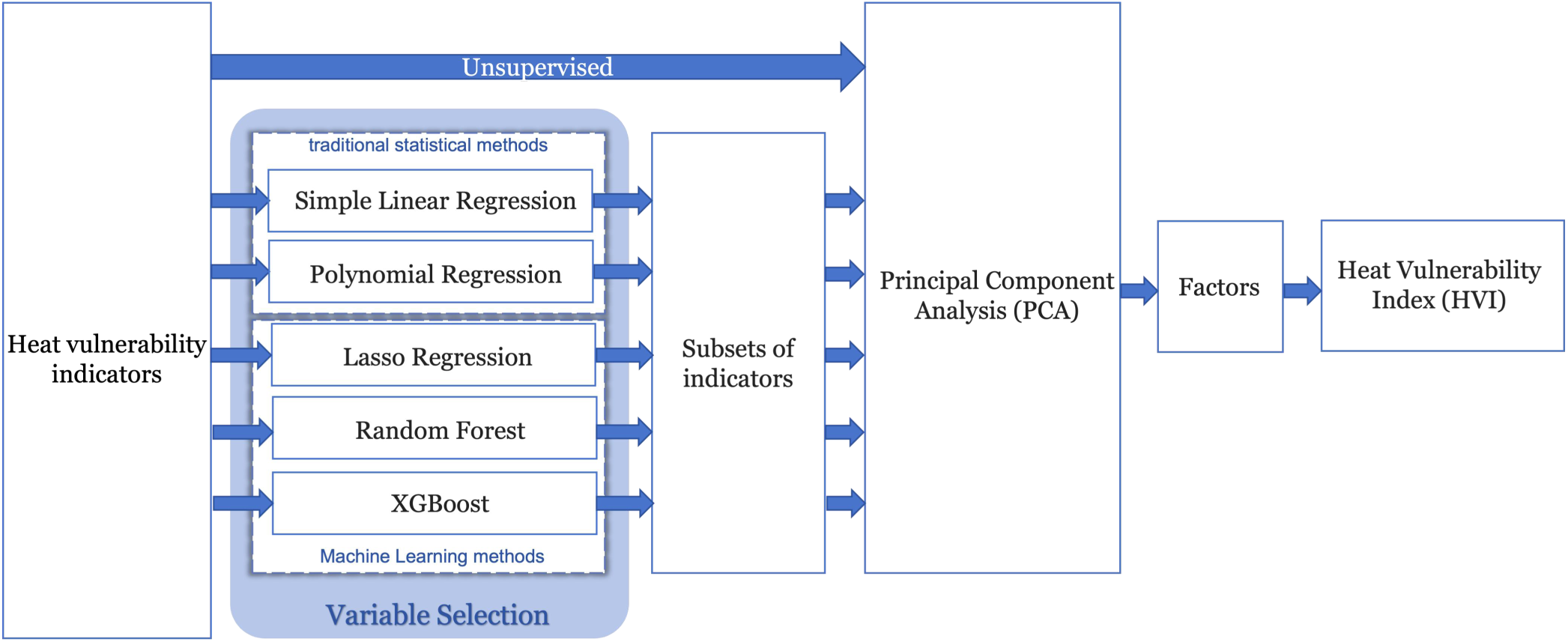
Study framework. Unsupervised Heat Vulnerability Index (HVI) models exclude data-driven variable selection, whereas supervised HVIs incorporate it. Heat vulnerability indicators serve as candidate predictors, and heat-related excess mortality serves as the outcome.

#### 3.4.1 SLR and PR

We first fitted univariate simple linear regression (SLR) models using ordinary least squares (OLS) to identify initial linear associations between each candidate indicator and heat-related excess mortality. Indicators with statistically significant coefficients (p < 0.05) were retained. To capture potential nonlinear relationships, we further employed PR, an extension of SLR that incorporates polynomial terms (Hastie et al., 2009). We used a univariate PR model of degree 3 (including linear, quadratic, and cubic terms) to regress heat-related excess mortality on each indicator. A joint hypothesis test (F-test) was then used to assess the overall significance of the polynomial terms. Indicators with statistically significant joint F-tests (p < 0.05) were retained for HVI construction.

#### 3.4.2 Lasso

Lasso regression is a type of regularized linear regression that minimizes the sum of squared residuals with L1 regularization, which shrinks some coefficients and sets others to 0, thereby retaining important features (Tibshirani, 1996; Cai et al., 2018; Emmert-Streib & Dehmer, 2019; Karpinska et al., 2021; Lounici, 2008). Compared with OLS, Lasso can mitigate the effects of multicollinearity, reduce overfitting, and potentially improve the overall prediction accuracy and interpretability (Tibshirani, 1996; Muthukrishnan & Rohini, 2016; Emmert-Streib & Dehmer, 2019; Theng & Bhoyar, 2024). In this study, the dataset was randomly split into training and testing sets. The mean and standard deviation were calculated from the training set, and these parameters were then used to standardize both the training and testing sets. We employed LassoCV with 5-fold cross-validation on the training set to optimize the regularization parameter. Indicators with non-zero coefficients were selected for HVI construction.

#### 3.4.3 Random Forest

Random forest (RF) is an ensemble machine learning algorithm that can learn nonlinear relationships and interactions from data without explicit model specification (Grömping, 2009; Genuer et al., 2010; Speiser et al., 2019). It characterizes complex data structures by constructing multiple decision trees and aggregating their predictions (Breiman, 2001). RF provides variable importance measures that reflect the relative contribution of each feature to the fitted model (Nguyen et al., 2015). In this study, the RF consisted of 5,000 decision trees, and 5 features were randomly selected at each tree split. The minimum number of split samples was set to 5 to control the complexity and avoid overfitting. Feature importance was evaluated using the out-of-bag (OOB) permutation importance, and the top 50% most important indicators were retained for subsequent analysis, which corresponded to an importance threshold of 0.01.

#### 3.4.4 XGBoost

XGBoost is a scalable machine learning method based on gradient boosting decision trees (Chen et al., 2016). In boosted tree models, gain is a primary metric used to quantify the importance of a feature (Zheng et al., 2017). The average gain is the total gain of all trees divided by the total number of splits in which the feature is used (Shi et al., 2019). Higher feature importance scores indicate greater contribution of a feature during model training (Shi et al., 2019; Jiang et al., 2023). We employed the built-in, average gain-based feature importance metric of XGBoost to rank the candidate indicators according to their relative contributions to predicting heat-related excess mortality. The dataset was randomly split into training and testing sets. The hyperparameter settings of the model were determined using a 5-fold cross-validation on the training set. By setting the cumulative importance threshold at 80%, a subset of indicators was selected to retain the most influential indicators while reducing dimensionality. This procedure utilizes XGBoost’s capacity to capture nonlinear relationships and feature interactions.

### 3.5 Evaluation and Sensitivity Analysis

To validate the resulting HVIs, we first treated both HVI scores and heat-related excess mortality as continuous variables. Spearman’s correlation coefficient was used to test the monotonic association between them. The mean squared error (MSE) and mean absolute error (MAE) were also calculated.

We then converted both measures into categorical variables using equal width binning, divided them into low, medium-low, medium-high, and high heat vulnerability categories, and assigned integer scores of 1 to 4 to these categories accordingly. To assess the ability of HVIs to differentiate communities across heat vulnerability categories, accuracy and F1 scores were employed as classification performance metrics.

To further evaluate the robustness of key heat vulnerability indicators, we conducted sensitivity analyses under different analytical settings: (1) adoption of a different definition of heat days, using the daily maximum HI with a threshold of 110 °F (43.3 °C); and (2) performance of age standardization on heat-related excess mortality based on the 2000 U.S. Standard Population, calculating weighted heat-related deaths by age group to reduce biases from population age structure differences across CAs. These procedures were systematically applied across all HVI approaches, resulting in 20 analytical scenarios (with/without age standardization × 2 heat day definitions × 5 variable selection approaches). Across these scenarios, we assessed the consistency of selected indicators to examine the robustness of the selection results.

In addition, potential lag effects of heat-related deaths over the study period were also assessed. We included the average daily deaths on days 1, 2, and 3 following each heat day and constructed scenarios with different lag periods (lag = 1, 2, and 3 days) for comparison. The results showed no significant differences whether the lag effect was considered or not (results not shown).

This study was approved by the Northwestern University Institutional Review Board (STU00219292).

## 4. Results

### 4.1 Heat-Related Excess Mortality

We analyzed the temporal trend of heat days (at least two consecutive days with the daily minimum HI exceeding 70 °F (21.1 °C) over the 27-year period via linear regression. The number of heat days over time shows an upward trend, with a regression slope of 0.09 heat days per year; however, the trend is not statistically significant (p = 0.61). Peak heat day occurrence was in 1995 (27 days) (Figure 2).

**Figure 2.**
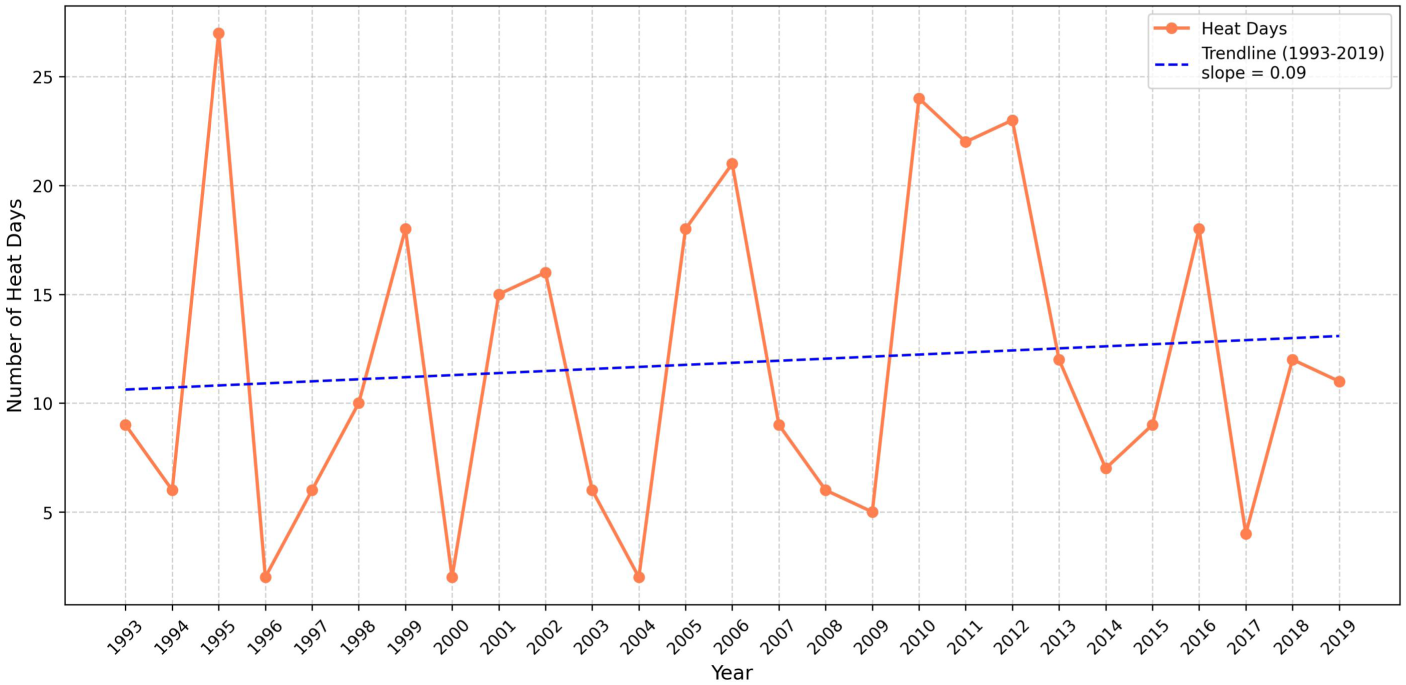
Annual number of heat days and linear trend (slope = 0.09, p = 0.61) in Chicago, 1993-2019. Heat days are defined as at least two consecutive days with the daily minimum Heat Index exceeding 70°F (21.1°C).

During this period, the average heat-related excess mortality in Chicago was ∼48 deaths per year, with ∼ 3 heat-related deaths per heat day (Figure 3). Significant peaks were observed in 1995 and 1999. In 1995, 675 heat-related deaths are estimated, with an average of 25.0 heat-related deaths per heat day; in 1999, there were 136 heat-related deaths and an average of 7.6 heat-related deaths per heat day. Both annual and daily heat-related excess mortality exhibit an overall declining pattern over this period (Figure 3), as illustrated by the linear trend lines shown in Supplemental Figure S3. To assess whether this long-term trend is disproportionately influenced by the extreme 1995 heatwave in Chicago, we additionally present the time series excluding 1995 in Supplemental Figure S4–5, which show consistent declining trends.

**Figure 3.**
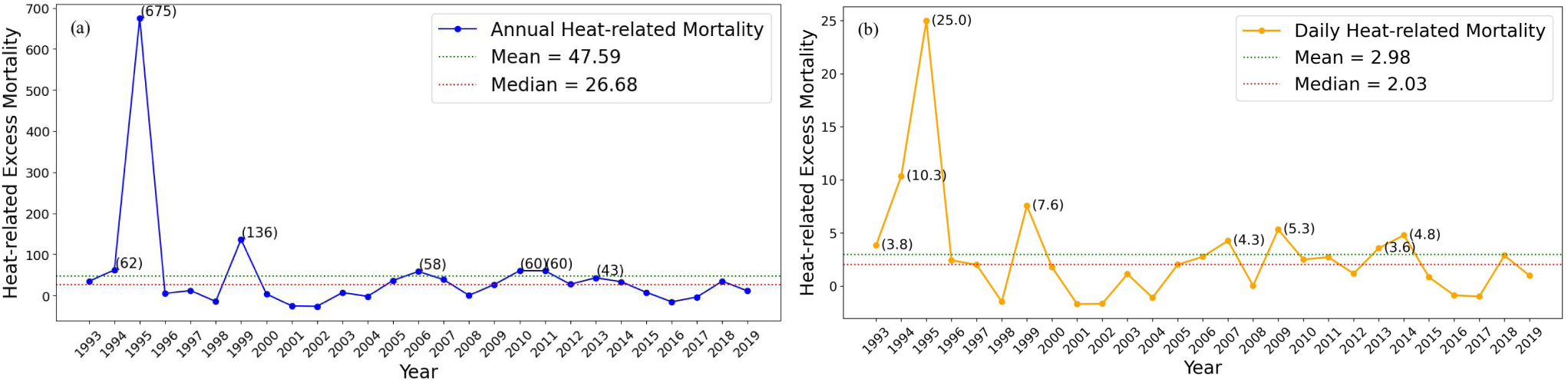
Time series of heat-related excess mortality in Chicago, 1993-2019. (a) Annual heat-related excess mortality; (b) Average daily heat-related excess mortality. Mean and median reference lines are shown, and values exceeding the mean are annotated.

The spatial distribution of annual and daily average heat-related excess mortality rate across the 77 CAs in Chicago shows that higher values are predominantly concentrated in the southern and western parts of the city (Figure 4). The Moran’s I values (0.19 for annual and 0.17 for daily, with both p < 0.05) indicate modest but statistically significant positive spatial autocorrelation, suggesting that communities with similar levels of heat-related health risk are often found clustered near one another.

**Figure 4.**
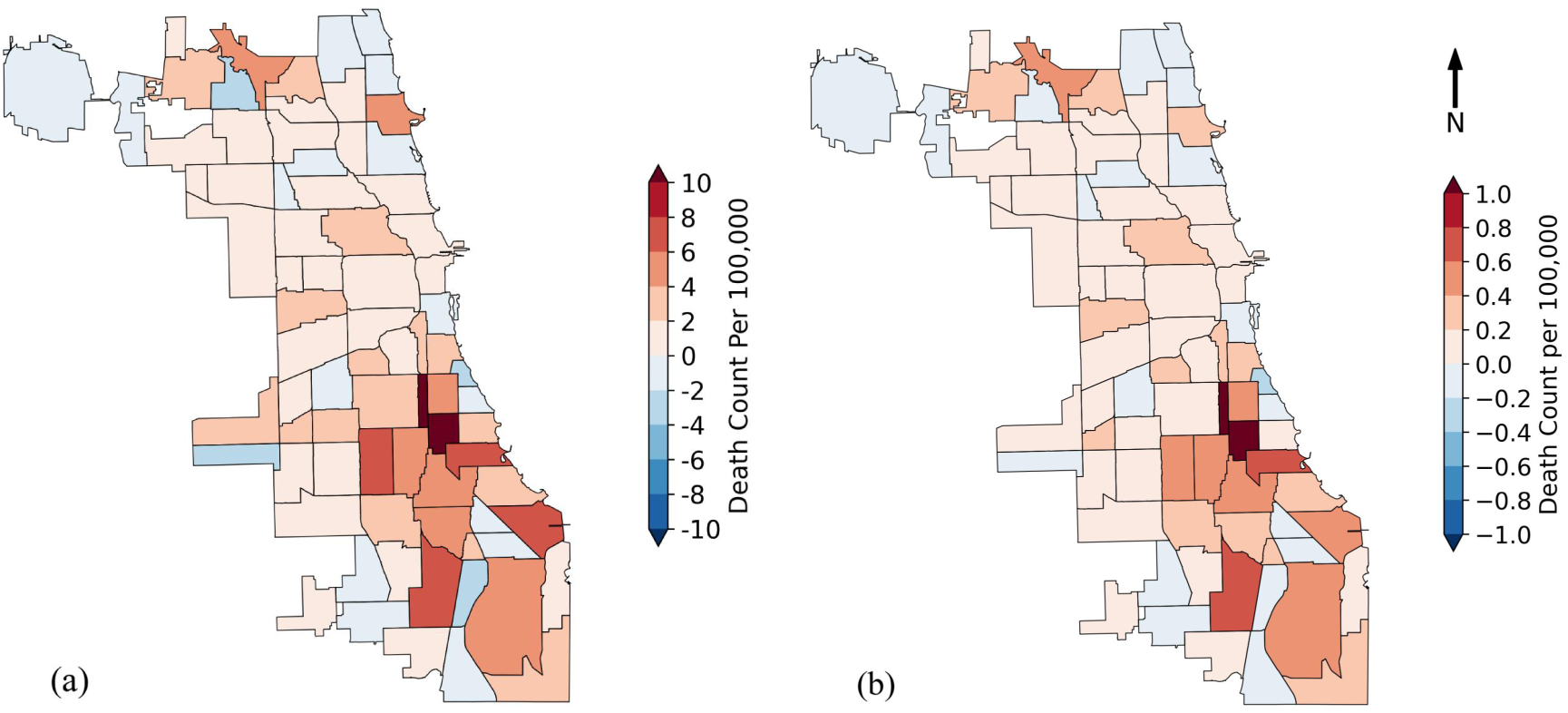
Spatial distribution of heat-related excess mortality rate across communities in Chicago, 1993–2019. Heat days are defined as at least two consecutive days with the daily minimum Heat Index exceeding 70°F (21.1°C). (a) Map of the average annual heat-related excess mortality rate (range:-2.6 to 13.2 per 100,000; mean: 1.8 per 100,000). Spatial autocorrelation is statistically significant (Moran’s I = 0.194, p = 0.008). (b) Map of the average daily heat-related excess mortality (range:-0.22 to 1.11 per 100,000; mean: 0.16 per 100,000). Spatial autocorrelation is statistically significant (Moran’s I = 0.168, p = 0.009).

### 4.2 Variable Selection

The heat vulnerability indicators used in the unsupervised HVI, as well as the indicators identified by various selection methods are summarized in Table 3. Indicators selected by each method are highlighted with a blue background. Detailed results for each method are provided in the Supplemental Materials (Supplemental Tables S2-S5 and Supplemental Figure S6). Results show that *PovertyRate*is a key indicator in all variable selection methods, while *NoACAccess*and *Ageabove65*remain important indicators in four out of the five variable selection methods. *LivingAlone*and *HispanicorLatino(ofAny Race)*were not selected by any of the methods, indicating a lack of robust association with heat-related excess mortality in Chicago.

**Table 3.** Variable selection and Principal Component Analysis results. Indicators used to construct the unsupervised HVI and those retained by each variable selection method for the supervised HVIs (SLR, PR, Lasso, RF, and XGBoost) are highlighted with a blue background. Factor loadings for heat vulnerability indicators are presented along with eigenvalues and the proportion of variance explained by each factor. Absolute values > 0.4 are considered the most significant loadings on that factor and are marked in bold.

| Indicator | Unsupervised HVI |  |  |  | SLR |  |  | PR |  |  | LASSO |  |  |  | RF |  |  | XGBoost |  |  |
| --- | --- | --- | --- | --- | --- | --- | --- | --- | --- | --- | --- | --- | --- | --- | --- | --- | --- | --- | --- | --- |
|  | Factor1 | Factor2 | Factor3 | Factor4 | Factor1 | Factor2 | Factor3 | Factor1 | Factor2 | Factor3 | Factor1 | Factor2 | Factor3 | Factor4 | Factor1 | Factor2 | Factor3 | Factor1 | Factor2 | Factor3 |
| Not Green Space | -0.05 | -0.01 | 0.02 | **0.91** |  |  |  |  |  |  | 0.00 | 0.01 | **1.00** | 0.00 |  |  |  | 0.00 | -0.01 | **0.94** |
| Age above 65 | -0.07 | -0.15 | **0.74** | -0.11 | 0.01 | **0.97** | -0.01 | 0.00 | 0.04 | **0.96** | 0.00 | **0.91** | 0.01 | 0.06 | 0.02 | -0.01 | **0.99** | 0.03 | **0.69** | -0.18 |
| Age above 65 and Living Alone | 0.24 | -0.35 | -0.02 | 0.21 | 0.08 | 0.00 | **0.91** | 0.29 | **-0.58** | -0.10 |  |  |  |  | 0.39 | **-0.63** | -0.02 |  |  |  |
| Living Alone | 0.02 | **-0.51** | -0.01 | 0.20 |  |  |  |  |  |  |  |  |  |  |  |  |  |  |  |  |
| Black (Non-Hispanic) | **0.47** | -0.15 | 0.15 | -0.09 | **0.51** | 0.20 | 0.10 | **0.46** | -0.22 | 0.23 |  |  |  |  |  |  |  | **0.57** | 0.13 | -0.06 |
| Hispanic or Latino (of Any Race) | -0.09 | **0.51** | -0.05 | 0.16 |  |  |  |  |  |  |  |  |  |  |  |  |  |  |  |  |
| Diabetes Prevalence | 0.07 | 0.18 | **0.63** | 0.12 |  |  |  |  |  |  | **0.70** | 0.30 | -0.02 | -0.17 |  |  |  | -0.01 | **0.70** | 0.18 |
| Less than High School Diploma | 0.27 | **0.53** | 0.08 | 0.10 |  |  |  | 0.30 | **0.78** | -0.06 | **0.71** | -0.29 | 0.02 | 0.17 | 0.31 | **0.77** | -0.02 |  |  |  |
| Poverty Rate | **0.63** | 0.04 | -0.14 | -0.09 | **0.54** | -0.14 | 0.20 | **0.59** | -0.01 | -0.13 | 0.00 | 0.05 | 0.00 | **0.97** | **0.65** | -0.03 | -0.08 | **0.63** | -0.13 | -0.09 |
| No AC Access | **0.49** | 0.05 | 0.00 | 0.12 | **0.67** | -0.05 | -0.35 | **0.51** | 0.09 | 0.04 |  |  |  |  | **0.57** | 0.05 | 0.09 | **0.52** | 0.00 | 0.19 |
| Eigenvalue | 3.55 | 2.29 | 1.27 | 0.95 | 2.75 | 0.99 | 0.71 | 2.76 | 1.29 | 0.98 | 1.66 | 1.19 | 0.91 | 0.79 | 2.06 | 1.26 | 0.97 | 2.53 | 1.23 | 0.97 |
| % Variance explained | 35.0 | 22.6 | 12.6 | 9.4 | 54.3 | 19.6 | 14.0 | 45.4 | 21.2 | 16.2 | 32.8 | 23.6 | 18.0 | 15.6 | 40.6 | 24.9 | 19.2 | 41.6 | 20.2 | 16.0 |

### 4.3 PCA

We conducted PCA on two sets of indicators: (1) the full set of candidate heat vulnerability indicators to construct an unsupervised HVI, and (2) separately on each subset of indicators selected by the five supervised methods (SLR, PR, Lasso, RF, XGBoost). The factor loadings, eigenvalues, and proportion of variance explained for all six PCA models are presented in Table 3.

For the unsupervised HVI, four factors were retained. Factor 1, which explains the maximum variance (35%), is comprised of *Poverty Rate*, *No AC Access*, and *Black (Non-Hispanic)*. Factor 2 (22.6% of variance) is characterized by a positive loading for *Less than High School Diploma* and *Hispanic or Latino (of Any Race)*, and a negative loading for *Living Alone*. Factor 3 (12.6% of variance) shows significant positive loadings for *Age above 65* and *Diabetes Prevalence*. Factor 4 (9.4% of variance) is dominated by *Not Green Space*.

PCA on the indicators selected by the SLR method identified three factors. Factor 1 (54.3% of variance) is comprised of *Poverty Rate*, *No AC Access*, and *Black (Non-Hispanic)*. *Age above 65* and *Age above 65 and Living Alone* form separate factors, explaining 19.6% and 14.0% of the variance, respectively.

PCA on the indicators selected by the PR method also identified three factors. Factor 1 explains 45.4% of the variance and consists of three indicators: *Poverty Rate*, *No AC Access*, and *Black (Non-Hispanic)*. *Less than High School Diploma* and *Age above 65 and Living Alone* together constitute Factor 2, explaining 21.2% of the variance. Notably, these two indicators show opposing loadings on this factor, indicating that their contributions to this factor are in opposite directions, which suggests that communities with a higher proportion of residents lacking a high school diploma tend to have a lower prevalence of adults aged over 65 living alone. Factor 3 (16.2% of the variance) is dominated by *Age above 65*.

PCA on the indicators selected by the Lasso method identified four factors. Factor 1, consisting of *Less than High School Diploma* and *Diabetes Prevalence*, explains 32.8% of the variance. The other three factors are dominated by *Age above 65*, *Not Green Space* and *Poverty Rate*, explaining 23.6%, 18.0% and 15.6% of the variance, respectively.

PCA on the indicators selected by the RF method extracted three factors. Factor 1 consists of *Poverty Rate* and *No AC Access*, which explains 40.6% of the variance. Factor 2 is similar to the PR result, characterized by opposite loadings between *Less than High School Diploma* and *Age above 65 and Living Alone*. Factor 3 is largely represented by *Age above 65*.

PCA on the indicators selected by the XGBoost method identified three factors. Factor 1, composed of *Poverty Rate*, *No AC Access* and *Black (Non-Hispanic)*, explains 41.6% of the variance. Factor 2 loads highly on *Age above 65* and *Diabetes Prevalence* and explains 20.2% of the variance. Factor 3 is dominated by *Not Green Space*, explaining 16.0% of the variance.

Overall, certain indicators consistently load strongly on the main factors. The first principal components (Factor 1), which explain maximum variance, always include *Poverty Rate* and *No AC Access*, and sometimes also include *Black (Non-Hispanic)*. The only exception occurs in the PCA results using indicators selected by Lasso, where neither *Poverty Rate* nor *No AC Access* appears in Factor 1, but *Poverty Rate* loads separately on Factor 4, explaining 16% of the variance. *Age above 65* is also a robust and important indicator, loading significantly on at least one factor in all methods. In addition, *Not Green Space* often forms a separate single factor, indicating that as an environmental variable, it is less related to the other heat vulnerability indicators and represents an independent dimension of heat vulnerability.

### 4.4 Construction and Evaluation of HVIs

HVIs constructed from different variable selection methods have distinct spatial patterns but predominantly identify the southern and western community areas (CAs) as those that are most vulnerable to heat (Figure 6). The unsupervised HVI (Figure 6a) shows that areas with higher vulnerability are mainly concentrated in the southern and western CAs. The SLR model produces HVI scores with higher values mainly concentrated in the southeastern CAs (Figure 6b). The PR-informed HVI (Figure 6c) indicates higher heat vulnerability in the southern and western CAs. The Lasso model generates HVI scores (Figure 6d) showing a general trend of higher vulnerability in the west and south. The RF model has the highest HVI score values concentrated in the southern region (Figure 6e), while the XGBoost model yields HVI highest in the west and south (Figure 6f).

**Figure 6.**
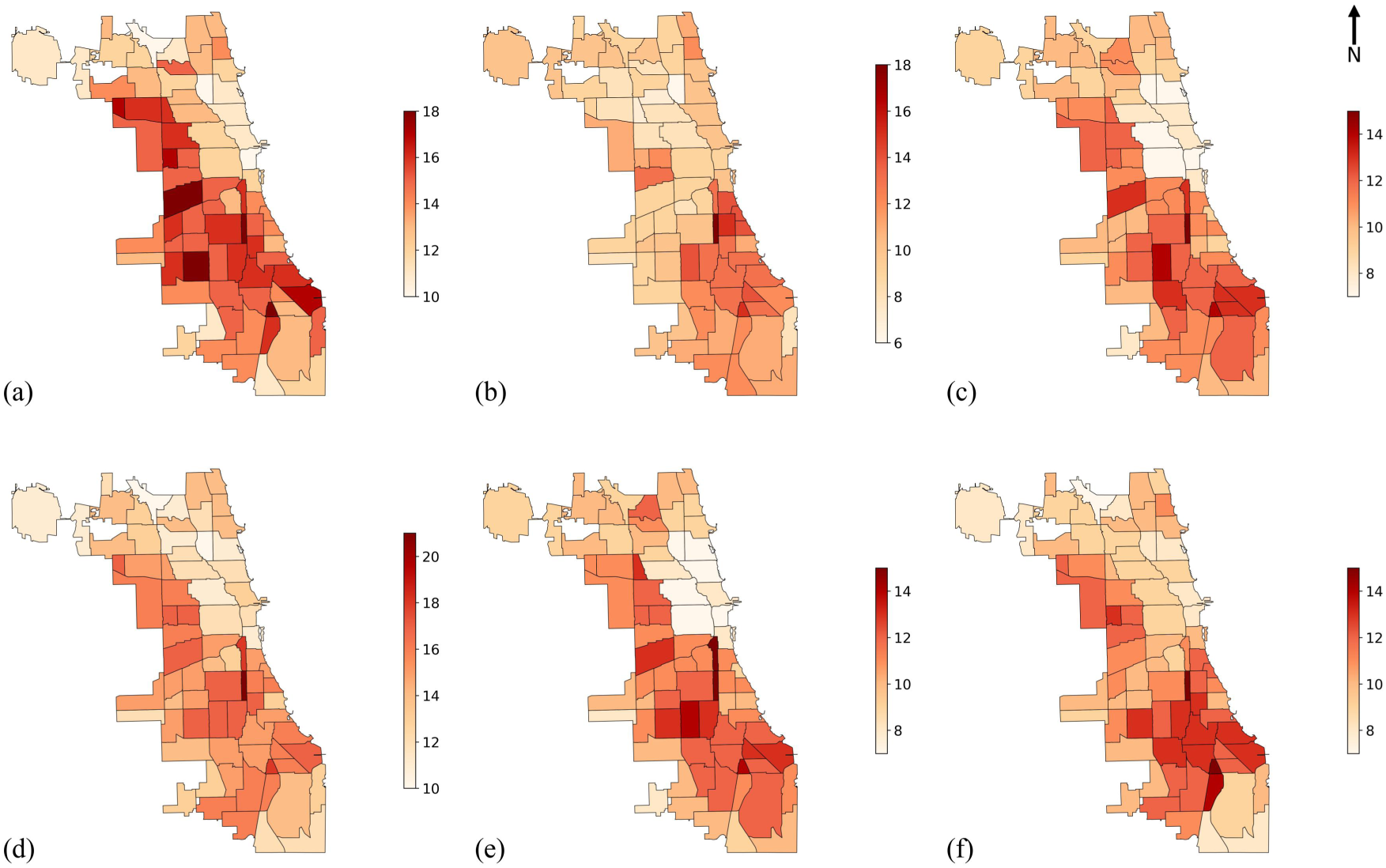
Heat Vulnerability Index (HVI) maps. HVI scores were generated using indicator subsets selected by different methods: (a) Unsupervised HVI; (b) Simple Linear Regression; (c) Polynomial Regression; (d) Lasso Regression; (e) Random Forest; (f) XGBoost. Higher HVI scores indicate higher heat vulnerability.

Since each PCA was performed on a different subset of indicators, the variance structure explained and the number of retained factors differ. As a result, the absolute HVI scores are on different numerical scales and cannot be directly compared across methods. To enable cross-method comparison, we transformed the HVIs to a unified ordinal scale by categorizing scores into quartiles (Levels 1–4). Figure 7 presents the spatial distribution of resulting heat vulnerability levels. Both the unsupervised HVI (Figure 7a) and the XGBoost-informed HVI (Figure 7f) identify scattered high vulnerability CAs (Level 4). The other methods identify fewer and more spatially clustered high heat vulnerability areas, with overlap among models in several southern CAs (Figure 7b–e). For the medium-high heat vulnerability areas (Level 3), most maps show a band-like distribution extending from the northwest to the southeast, with the exception of the SLR-informed method, which exhibits higher heat vulnerability in eastern CAs (Figure 7b). For medium-low heat vulnerability areas (Level 2), the distribution generally surrounds high and medium-high vulnerability CAs, without obvious spatial clustering. Low vulnerability areas (Level 1) are relatively uniformly distributed, primarily along the northern and southern margins of the city, as well as in several CAs near Lake Michigan in the northeast. In summary, despite variations in the precise location and extent of the highest-risk clusters, HVI maps converge on a common pattern in which Chicago’s northeastern CAs generally exhibit lower heat vulnerability, while higher vulnerability areas are mostly concentrated in the southern and western parts of the city.

**Figure 7.**
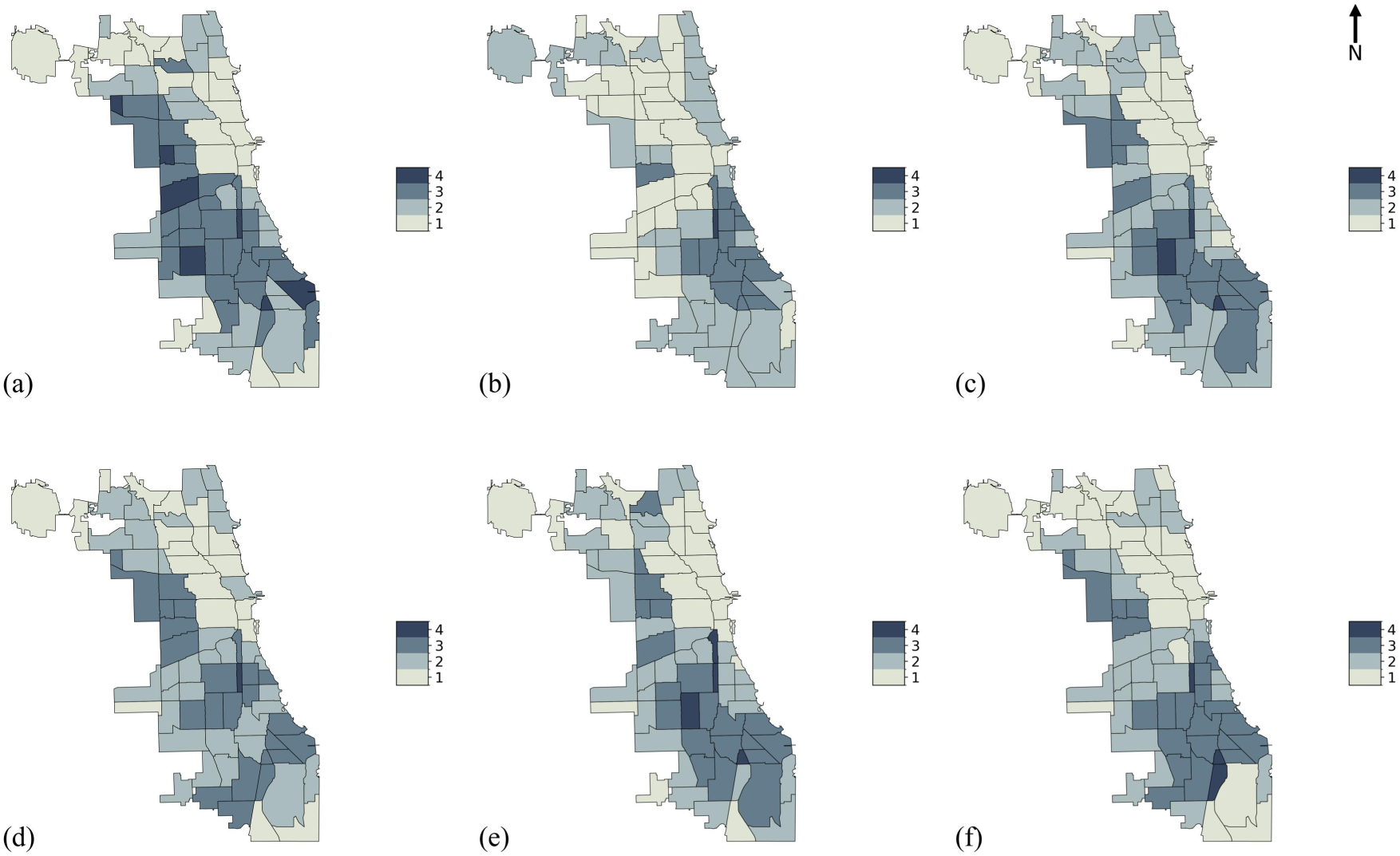
Heat Vulnerability Index (HVI) category maps. HVI scores were generated using indicator subsets selected by different methods: (a) Unsupervised HVI; (b) Simple Linear Regression; (c) Polynomial Regression; (d) Lasso Regression; (e) Random Forest; (f) XGBoost. The HVI scores were classified into four categories using equal width binning: 1 = Low, 2 = Medium-low, 3 = Medium-high, 4 = High. Higher levels indicate higher heat vulnerability.

To evaluate the performance of each HVI, we assess the correlation and categorical classification metrics between HVI scores and estimated heat-related excess mortality for each of the 77 CAs. The results of both continuous evaluation metrics (Spearman correlation, MAE, and MSE) and categorical classification metrics (accuracy and F1 score) are summarized in Table 4. For continuous evaluation, the RF-informed HVI exhibited the highest Spearman correlation coefficient with heat-related excess mortality (ρ = 0.37), exceeding that of the unsupervised HVI (ρ = 0.29) and the SLR-informed HVI (ρ = 0.28). This level of correlation suggests a moderate monotonic association. In addition, compared with the other HVIs, the RF-informed HVI demonstrated relatively lower MAE and MSE values, indicating its superior overall performance under continuous evaluation metrics. In categorical evaluation, the RF-informed HVI achieves an accuracy of 0.49 and F1 score of 0.51, both of which are the best among the evaluated HVIs. These values are 0.17 and 0.16 higher than those of the unsupervised HVI (Accuracy = 0.32, F1 score = 0.35), corresponding to relative increases of approximately 53% and 46%.

**Table 4.** Performance metrics of different Heat Vulnerability Index models for continuous and categorical tasks. Spearman’s ρ: Spearman’s rank correlation coefficient; higher values indicate stronger monotonic agreement between predicted heat vulnerability and observed heat-related excess mortality. Mean Absolute Error (MAE) and Mean Squared Error (MSE): Lower values indicate better model performance. Accuracy: Proportion of correct classifications in categorical tasks; higher values indicate better performance. F1 score: Harmonic mean of precision and recall in categorical tasks; higher values indicate better balance between precision and recall.

| HVI Models | Spearman’s $\rho$<br>(Continuous) | MAE<br>(Continuous) | MSE<br>(Continuous) | Accuracy<br>(Categorical) | F1 score<br>(Categorical) |
| --- | --- | --- | --- | --- | --- |
| Unsupervised HVI | 0.29 | 13.87 | 196.13 | 0.32 | 0.35 |
| SLR | 0.28 | 10.41 | 112.92 | 0.42 | 0.42 |
| PR | 0.32 | 10.39 | 110.69 | 0.47 | 0.48 |
| LASSO | 0.30 | 14.23 | 206.79 | 0.45 | 0.48 |
| RF | 0.37 | 10.41 | 111.61 | 0.49 | 0.51 |
| XGBoost | 0.28 | 10.45 | 112.08 | 0.45 | 0.46 |

The PR-informed HVI achieves the lowest MAE and MSE among all models, while its Spearman correlation and classification accuracy are slightly lower than those of the RF-informed HVI. In continuous prediction, the HVIs informed by SLR and XGBoost have the poorest Spearman correlation values among methods despite having lower MAE and MSE, whereas the Lasso-informed HVI produces the highest MAE and MSE. In categorical prediction, the SLR-informed HVI shows an improvement over the unsupervised HVI but remains inferior to both RF-and PR-informed HVIs (Table 4).

### 4.5 Sensitivity Analyses

To examine the robustness of several of our assumptions, we conducted sensitivity analyses using different analytical settings, including: (1) an alternative heat day definition based on the daily maximum Heat Index (HI >110°F (43.3°C)), with corresponding heat-related excess mortality maps shown in Supplemental Figure S7; (2) age-standardized heat-related mortality data; and (3) lag effects of heat-related deaths.

These analyses suggest that the key heat vulnerability indicators are robust across analytical settings. That is, regardless of extreme heat definition or age standardization, *Poverty Rate, No AC Access*, and *Age above 65* are consistently identified as important indicators by most variable selection methods, while *Less than High School Diploma*, and *Black (Non-Hispanic)* also show higher selection frequency compared with the other indicators (Figure 8).

**Figure 8.**
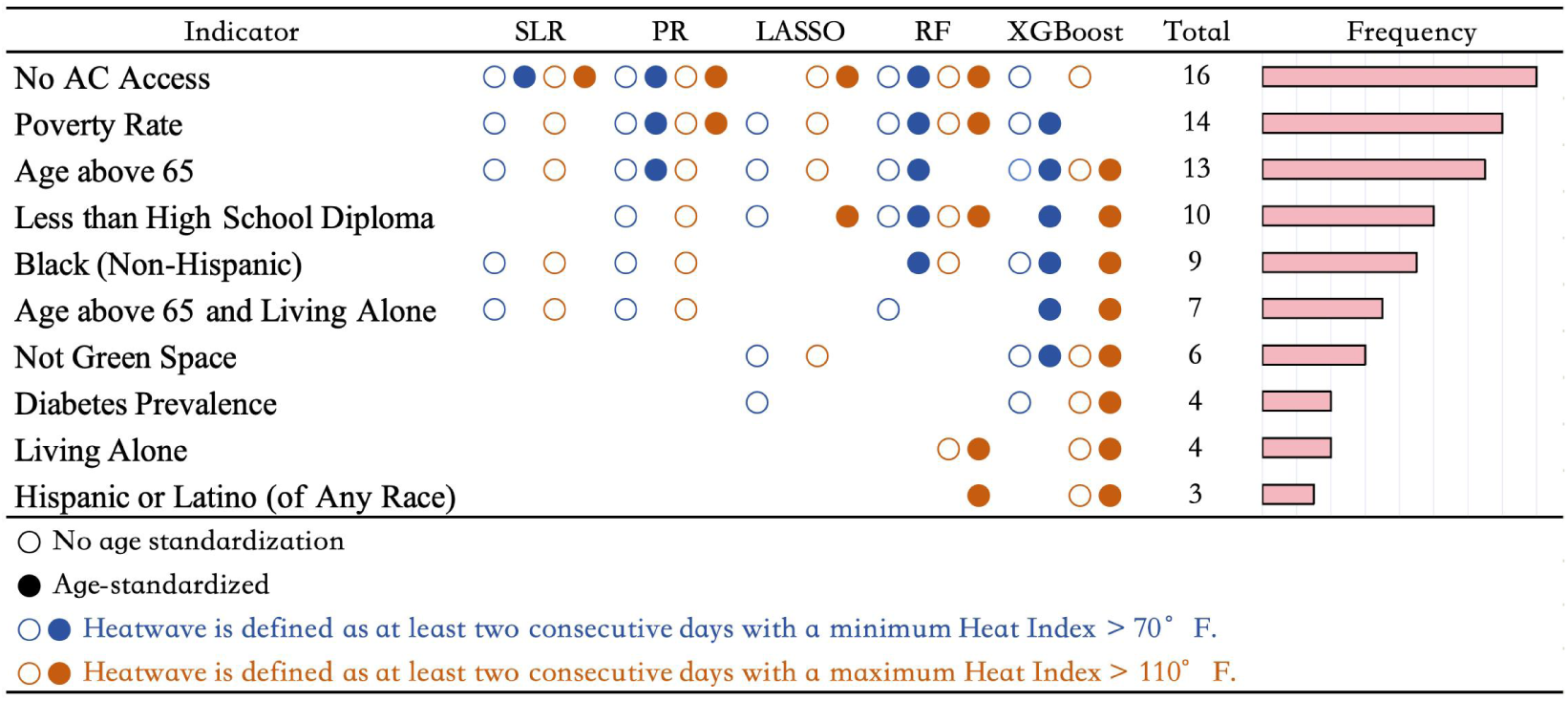
Variable selection results across different analytical configurations. Two configurations were considered: (1) weather indicator defined as daily maximum Heat Index > 110°F (43.3°C) or daily minimum Heat Index > 70°F (21.1°C), and (2) age-standardized vs. non-standardized heat-related mortality. The bar plot shows the selection count of each indicator, with indicators ordered by selection frequency.

## 5. Discussion

This study systematically compares the performance of various variable selection methods in the construction of PCA-based HVIs, using a set of established heat vulnerability indicators derived from the published literature. The results indicate that, under our study design and data conditions, *No AC Access, Poverty Rate,* and *Age above 65* at the community level are the most robust determinants of heat vulnerability in Chicago. Random Forest-based variable selection outperforms other methods when the results are evaluated against heat-related excess mortality data, suggesting that supervised machine learning-based variable selection holds great promise for improving heat vulnerability assessments.

The identification of *Poverty Rate* and *No AC Access* in our analysis aligns with previous retrospective analyses of Chicago heatwaves. During the 1995 Chicago heatwave, deaths were more prevalent in the low-income areas of the city, and neighborhood affluence was negatively associated with heatwave mortality (Klinenberg, 1999; Browning et al., 2006). In both the 1995 and 1999 heatwaves, access to AC was consistently associated with lower death risk and was identified as a strong protective factor (Semenza et al., 1996; Naughton et al., 2002). Our results also align with broader global patterns. Existing empirical studies in multiple countries and cities have shown that, under heat exposure, economically disadvantaged populations often exhibit higher mortality rates or disease burdens (de Visser et al., 2022; Lim et al., 2015; Toloo et al., 2014; Fletcher et al., 2012; Madrigano et al., 2013; Chan et al., 2012; Zanobetti et al., 2013; Curriero et al., 2002; Greenberg et al., 1983), while an increase in AC prevalence has been linked to lower heat-related mortality and hospitalization rates (Anderson & Bell, 2009; Curriero et al., 2002; O’Neill et al., 2005; Greenberg et al., 1983; Ostro et al., 2010; Bouchama et al., 2007; Davis et al., 2003).

In addition to the vulnerability indicators discussed above, older age, race, and educational attainment were also frequently identified as important indicators in this study. During the 1995 Chicago heatwave, Black residents and older adults faced a disproportionately higher risk of death (Kaiser et al., 2007; Whitman et al., 1997; Healy, 2005; Klinenberg, 2022; Klinenberg, 1999; Naughton et al., 2002; Semenza et al., 1999). These early findings are further corroborated by more recent community level research on heat-related cardiovascular morbidity and mortality in Chicago, which identifies lower income, unemployment, lower college attainment, and a high proportion of Black residents as dominant contributors to structural vulnerability (Graffy et al., 2025). This pattern is also consistent with extensive epidemiological studies based on mortality rates or hospitalization outcomes, which suggest that older age, higher proportions of racial or ethnic minority populations, and lower educational attainment are significantly associated with higher heat-related health risks (Kephart et al., 2022; Kollanus et al., 2021; Graczyk et al., 2022; Thompson et al., 2022; Alahmad et al., 2020; Zhang et al., 2018; Yin et al., 2018; Kang et al., 2020; Fletcher et al., 2012; Stafoggia et al., 2006; Zanobetti et al., 2013; Lin et al., 2009; Medina-Ramón et al., 2006; O’Neill et al., 2003; Green et al., 2010; Uejio et al., 2011; Anderson & Bell, 2009; Michelozzi et al., 2005; Borrell et al., 2006; Curriero et al., 2002).

Some of our findings diverge from those reported in previous studies. For instance, *Living Alone* did not show consistent importance across various variable selection methods in our research. However, case-control studies reported that individuals living alone faced a higher risk of death during the 1995 and 1999 Chicago heatwaves (Semenza et al., 1996; Naughton et al., 2002). This difference may be related to the research scale. Klinenberg (1999) pointed out that this individual-level methodological approach leaves the differences between community areas invisible. Although living alone is an important individual risk factor, it may not serve as a robust predictor of heat-related mortality at the community level. Browning et al. (2006) found in a neighborhood-level quantitative analysis that while the proportion of residents living alone was significantly and positively associated with the baseline mortality, its explanatory power for the variation in mortality during the 1995 Chicago heatwave was not significant. Moreover, living alone is often interpreted as reflecting vulnerability arising from insufficient social support in heat vulnerability research, and conceptually is more regarded as a proxy variable for social isolation (Klinenberg, 1999). The strength of the social support network is not entirely determined by whether one is living alone. At the contemporary community scale in Chicago, its statistical effect may be overshadowed by more dominant structural factors such as poverty rate and lack of air conditioning. Therefore, the statistical importance of living alone may vary across research contexts and analysis scales.

In addition, the proportion of Hispanic or Latino population and the prevalence of diabetes have been linked to higher risk of heat-related mortality or morbidity in some research (Lin et al., 2009; Fletcher et al., 2012; Schwartz, 2005; Xu et al., 2019), but these indicators did not show robust importance across variable selection methods in our analysis. In contrast, we identified the proportion of Black residents as an important heat vulnerability indicator, whereas some prior studies did not find a significant association between race and certain heat-related health outcomes. Green et al. (2010) reported no significant effect modification by race on temperature-associated hospital admissions in nine counties in California; similarly, Pillai et al. (2014) found no significant differences between Whites and Black individuals in terms of the proportion of heat-related emergency department visits resulting in hospital admission versus routine discharge in Georgia. Shannon et al. (2025) found that extreme heat was associated with increased mortality among California veterans with cardiometabolic diseases, but reported no statistically significant effect modification by race. These authors attributed the absence of observed race effects primarily to differences in outcome definitions and institutional contexts. Specifically, hospital admissions may not fully capture heat-related morbidity across racial groups because of possible differences in health care utilization (Green et al., 2010), whereas the integrated Veterans Affairs care delivery paradigm may reduce racial disparities through targeted interventions addressing social factors related to health (Shannon et al., 2025). These examples collectively suggest that race does not always exhibit a stable association with heat-related health outcomes across settings, highlighting the strong context dependence of its statistical significance as a heat vulnerability indicator.

As regional contexts differ in population structure, health care systems, and health insurance coverage, heat vulnerability exhibits significant spatial heterogeneity. People in different regions respond differently to heat exposure (Gasparrini et al., 2015). Consequently, the key predictors for heat-related health outcomes can vary across regions (Anderson & Bell, 2009; Arsad et al., 2022). For example, a study by Mallen et al. (2019) in Dallas, Texas, found that older populations, people without a high school diploma, and lack of access to AC were important indicators of heat-related mortality. Whereas in Detroit, Michigan, Conlon et al. (2020) found that older age, socioeconomic status, race, and ethnicity indicators were not robust stand-alone determinants of heat vulnerability. In Phoenix, Arizona, Chuang & Gober (2015) found that lower socioeconomic status, the proportion of adults age >65 years living alone, the proportion of adults living alone, and the diabetes hospitalization rate could be used to predict heat vulnerability at the census tract level, while AC prevalence was not a significant predictor of heat-related health outcomes. Studies focused on non-U.S. locales show similar nuance. Li et al. (2025), in their study of heat vulnerability in Australian capital cities, suggested that personal health status and sociodemographic characteristics (including individual disease status, age, and education level) play a dominant role in determining heat vulnerability, overshadowing the influence of environmental and infrastructure factors. In contrast, Jeong et al. (2024), in a city-level study in South Korea, found that climate conditions and income among the elderly population showed statistically significant associations with heat-related mortality. These case studies from diverse locales indicate that the explanatory power of specific heat vulnerability indicators can be inconsistent across different geographic contexts, with each city exhibiting a distinct combination of important indicators.

These local nuances highlight that there is no single, stable set of heat vulnerability indicators that can be universally applied across different regional settings. Differences in climatic conditions, population structure, socioeconomic composition, and health care systems jointly shape distinct local heat-related health risk mechanisms across cities and regions. Given this context, our study emphasizes the importance of adopting supervised HVI frameworks where variable selection is informed by local heat-related health outcomes. Compared with unsupervised approaches, this data-driven step allows for the statistical identification of core combinations of indicators that are most explanatory of local risks, leading to more accurate heat vulnerability assessments that can better inform the spatial allocation of resources and targeted intervention strategies. Practically, this supports avoiding one-size-fits-all HVIs and instead selecting and validating indicators against local outcomes and community knowledge prior to using maps to focus interventions (Harris et al., 2018; Potash et al., 2020).

In this context, the choice of variable selection method can directly influence HVI outcomes, as different methods are grounded in distinct statistical assumptions and algorithmic mechanisms. Traditional SLR focuses on the linear relationship between the outcome variable (heat-related excess mortality) and each heat vulnerability indicator, and thus, less effectively captures potential nonlinear effects or complex interactions among indicators. In contrast, polynomial regression can fit a certain degree of nonlinear relationships by introducing higher order terms. In our variable selection results, PR identified the *Less than High School Diploma* indicator along with a set of indicators largely consistent with those identified by SLR. This result suggests a potential nonlinear association, where heat vulnerability may increase disproportionately once the proportion of residents with low educational attainment exceeds a certain level. However, PR is still based on a linear model framework with fixed parameter forms, which limits its ability to fully capture more complex nonlinear relationships and interactions.

As a machine learning approach, Lasso is also sensitive to linear relationships and tends to retain a representative variable in a highly correlated group while setting the coefficients of others to zero. This mechanism could lead to the omission of variables with practical significance. In this study, the AC access indicator was robustly identified by other methods, but not selected by Lasso. This is likely attributable to its moderate correlation with poverty rates (r = 0.59, see Supplemental Figure S2). Both RF and XGBoost are capable of capturing nonlinear effects. Although XGBoost is known for strong capabilities in capturing feature interactions and nonlinear signals, it underperformed across evaluation metrics in our study. This may not reflect a limitation of the XGBoost algorithm itself, but rather its heightened sensitivity to noise under the small sample size (n=77 CAs). The RF-informed HVI most accurately depicts the distribution of heat-related mortality, demonstrating the potential of this approach for developing precise heat vulnerability assessment tools under the conditions of this study. Overall, the performance of any variable selection method in this study should not be generalized across contexts. Rather than overinterpreting results from a single approach, greater emphasis should be placed on indicators that demonstrate stable selection across multiple methods. For implementation, these HVIs should be viewed as decision-support tools rather than sole eligibility screens. They are best used alongside local practitioner insight and lived experience and should be revisited as housing conditions, cooling access, demographics, and climate hazards change. Where feasible, cities should also pair HVI-based prioritization with monitoring of service uptake and health outcomes to reduce the risk of systematic under-identification of vulnerable groups.

While it is notable that our study identifies RF as the best performing supervised variable selection method, the resultant Spearman’s correlation coefficient (0.37) and classification accuracy of less than 50% leave ample room for improvement. One reason for the muted performance may be that we limited our candidate heat vulnerability indicators to those used by Reid et al. (2009). The choice was made intentionally to assess the HVI construction approach rather than to create a high-fidelity characterization of Chicago’s heat vulnerability. Since this candidate indicator set may not include certain key determinants of heat vulnerability specific to the Chicago context, the resulting HVIs are likely to only partially reflect the complexity of heat-related health risks in the city.

For higher-fidelity, location-specific applications, researchers should consider incorporating a comprehensive set of candidate indicators. The systematic review by Cheng et al. (2021) on heat vulnerability indices emphasizes the importance of including as many relevant indicators as possible. Alternatively, a community-informed approach to indicator identification could be used to select a broader set of locally-informed and contextually-relevant indicators; a methodological choice designed to leverage the knowledge of lived experience and to build trust amongst the community of users (Hay-Chapman et al., in prep).

Nevertheless, existing research has shown that HVIs can be sensitive to the scale, measurement, and the institutional and social context in which indicators are applied (Conlon et al., 2020; Chuang & Gober, 2015; Tate, 2012; Hinkel, 2011). Conlon et al. (2020) found that using different vegetation cover indicators resulted in HVI spatial distribution differences. Additionally, data limitations may constrain the explanatory power of the HVI. Limited temporal coverage of socioeconomic data often necessitates the assumption that vulnerability indicators change uniformly over time, i.e., that the relative differences between communities remain constant throughout the study period. Though this assumption is common in heat vulnerability research, it may lead to biased estimation, especially over longer study periods or when using temporally dynamic indicators.

## 6. Conclusion

This study compared the performance of unsupervised and supervised variable selection methods for the construction of heat vulnerability indices. Under the settings and data conditions of this study, a Random Forest-informed HVI exhibited the most robust performance, highlighting the potential of machine learning approaches to capture the nonlinear relationships and interactions between heat vulnerability indicators and heat-related health outcomes. This study provides empirical support for incorporating variable selection into HVI construction and underscores the importance of method choice. In heat vulnerability assessments, researchers should systematically evaluate variable selection strategies in accordance with specific research objectives, local context, and data characteristics. These findings suggest that neither indicator sets nor method performance can be assumed to be transferable across cities. Locally tailored, outcome-informed validation is therefore essential before HVI maps are used to inform equitable resource allocation for climate adaptation and heat risk mitigation.

## Data Availability

Meteorological data and socio-demographic data are publicly available from relevant data providers. Mortality data were obtained through institutional access and are not publicly available. Access to mortality data may be requested from the original data provider, subject to their approval.

https://www.census.gov/programs-surveys/acs.html

https://daymet.ornl.gov/

https://data.cityofchicago.org/

https://www.usgs.gov/centers/eros/science/national-land-cover-database

## Acknowledgment

This work was sponsored by the School of Integrated Climate and Earth System Sciences at University of Hamburg; the Office of Global Initiatives at the McCormick School of Engineering and the Paula M. Trienens Institute for Sustainability and Energy at Northwestern University; and the Buffett Institute for Global Affairs Defusing Disasters Working Group. We thank the Illinois and Chicago Departments of Public Health for providing the mortality records. JS, BP, LB and MvS acknowledge funding by the Deutsche Forschungsgemeinschaft (DFG, German Research Foundation) under Germany’s Excellence Strategy – EXC 2037 “CLICCS – Climate, Climatic Change, and Society” – Project Number 390683824.

## Author Contributions

Shuyue Qu: Conceptualization, Methodology, Software, Validation, Formal analysis, Investigation, Data curation, Writing – original draft, Writing – review & editing, Visualization.

Jana Sillmann: Conceptualization, Methodology, Resources, Writing – review & editing, Supervision, Project administration, Funding acquisition.

Benjamin W. Barrett: Conceptualization, Methodology, Software, Data curation, Writing – review & editing.

Peter M. Graffy: Conceptualization, Methodology, Software, Data curation, Writing – review & editing. Benjamin Poschlod: Methodology, Software, Writing – review & editing.

Lukas Brunner: Methodology, Writing – review & editing.

Raed Mansour: Conceptualization, Resources, Writing – review & editing. Malte von Szombathely: Writing – review & editing.

Finley Hay-Chapman: Writing – review & editing.

Teresa H. Horton: Conceptualization, Writing – review & editing. Jennifer Chan: Conceptualization, Writing – review & editing.

Sheetal Khedkar Rao: Conceptualization, Writing – review & editing. Kyra Woods: Conceptualization, Writing – review & editing.

Abel N Kho: Writing – review & editing, Supervision.

Daniel E. Horton: Conceptualization, Methodology, Resources, Writing – review & editing, Supervision, Project administration, Funding acquisition.

## References

1. Alahmad, B., Shakarchi, A. F., Khraishah, H., Alseaidan, M., Gasana, J., Al-Hemoud, A.,…& Fox, M. A. (2020). Extreme temperatures and mortality in Kuwait: Who is vulnerable?. Science of the Total Environment, 732, 139289.

2. Anderson, B. G., & Bell, M. L. (2009). Weather-related mortality: how heat, cold, and heat waves affect mortality in the United States. Epidemiology, 20(2), 205–213.

3. Arsad, F. S., Hod, R., Ahmad, N., Ismail, R., Mohamed, N., Baharom, M., Osman, Y., Radi, M. F. M., & Tangang, F. (2022). The Impact of Heatwaves on Mortality and Morbidity and the Associated Vulnerability Factors: A Systematic Review. International Journal of Environmental Research and Public Health, 19(23), 16356.

4. Azhar, G. S., Mavalankar, D., Nori-Sarma, A., Rajiva, A., Dutta, P., Jaiswal, A.,…& Hess, J. J. (2014). Heat-related mortality in India: excess all-cause mortality associated with the 2010 Ahmedabad heat wave. PloS one, 9(3), e91831.

5. Barnett, A. G., & Åström, C. (2012). Commentary: What measure of temperature is the best predictor of mortality?. Environmental Research, 118, 149–151.

6. Bell, M. L., Gasparrini, A., & Benjamin, G. C. (2024). Climate change, extreme heat, and health. New England Journal of Medicine, 390(19), 1793–1801.

7. Benmarhnia, T., Deguen, S., Kaufman, J. S., & Smargiassi, A. (2015). Vulnerability to heat-related mortality: A systematic review, meta-analysis, and meta-regression analysis. Epidemiology, 26(6), 781–793.

8. Borrell, C., Marí-Dell’Olmo, M., Rodríguez-Sanz, M., Garcia-Olalla, P., Caylà, J. A., Benach, J., & Muntaner, C. (2006). Socioeconomic position and excess mortality during the heat wave of 2003 in Barcelona. European journal of epidemiology, 21(9), 633–640.

9. Bouchama, A., Matthies, F., Shoukri, M., Dehbi, M., Mohamed, G., & Menne, B. (2007). Prognostic factors in heat wave–related deaths: A meta-analysis. Archives of Internal Medicine, 167(20), 2170–2176.

10. Breiman, L. (2001). Random forests. Machine learning, 45(1), 5–32.

11. Browning, C. R., Wallace, D., Feinberg, S. L., & Cagney, K. A. (2006). Neighborhood social processes, physical conditions, and disaster-related mortality: the case of the 1995 Chicago heat wave. American sociological review, 71(4), 661–678.

12. Cai, J., Luo, J., Wang, S., & Yang, S. (2018). Feature selection in machine learning: A new perspective. Neurocomputing, 300, 70–79.

13. Chan, E. Y. Y., Goggins, W. B., Kim, J. J., & Griffiths, S. M. (2012). A study of intracity variation of temperature-related mortality and socioeconomic status among the Chinese population in Hong Kong. J Epidemiol Community Health, 66(4), 322–327.

14. Chen, T., & Guestrin, C. (2016, August). Xgboost: A scalable tree boosting system. In Proceedings of the 22nd acm sigkdd international conference on knowledge discovery and data mining (pp. 785-794).

15. Cheng, W., Li, D., Liu, Z., & Brown, R. D. (2021). Approaches for identifying heat-vulnerable populations and locations: A systematic review. Science of The Total Environment, 799, 149417.

16. Chuang, W. C., & Gober, P. (2015). Predicting hospitalization for heat-related illness at the census-tract level: accuracy of a generic heat vulnerability index in Phoenix, Arizona (USA). Environmental health perspectives, 123(6), 606–612.

17. Conlon, K. C., Mallen, E., Gronlund, C. J., Berrocal, V. J., Larsen, L., & O’Neill, M. S. (2020). Mapping human vulnerability to extreme heat: A critical assessment of heat vulnerability indices created using principal components analysis. Environmental health perspectives, 128(9), 097001.

18. Curriero, F. C., Heiner, K. S., Samet, J. M., Zeger, S. L., Strug, L., & Patz, J. A. (2002). Temperature and mortality in 11 cities of the eastern United States. American journal of epidemiology, 155(1), 80–87.

19. Davis, R. E., Knappenberger, P. C., Michaels, P. J., & Novicoff, W. M. (2003). Changing heat-related mortality in the United States. Environmental health perspectives, 111(14), 1712.

20. de Visser, M., Kunst, A. E., & Fleischmann, M. (2022). Geographic and socioeconomic differences in heat-related mortality among the Dutch population: a time series analysis. BMJ open, 12(11), e058185.

21. Ebi, K. L., Capon, A., Berry, P., Broderick, C., de Dear, R., Havenith, G.,…& Jay, O. (2021). Hot weather and heat extremes: health risks. The lancet, 398(10301), 698–708.

22. Emmert-Streib, F., & Dehmer, M. (2019). High-dimensional LASSO-based computational regression models: regularization, shrinkage, and selection. Machine Learning and Knowledge Extraction, 1(1), 359–383.

23. Fletcher, B. A., Lin, S., Fitzgerald, E. F., & Hwang, S. A. (2012). Association of summer temperatures with hospital admissions for renal diseases in New York State: a case-crossover study. American journal of epidemiology, 175(9), 907–916.

24. Fouillet, A., Rey, G., Laurent, F., Pavillon, G., Bellec, S., Guihenneuc-Jouyaux, C.,…& Hémon, D. (2006). Excess mortality related to the August 2003 heat wave in France. International archives of occupational and environmental health, 80(1), 16–24.

25. Gasparrini, A., Guo, Y., Hashizume, M., Lavigne, E., Zanobetti, A., Schwartz, J.,…& Armstrong, B. (2015). Mortality risk attributable to high and low ambient temperature: a multicountry observational study. The lancet, 386(9991), 369–375.

26. Genuer, R., Poggi, J. M., & Tuleau-Malot, C. (2010). Variable selection using random forests. Pattern recognition letters, 31(14), 2225–2236.

27. Graczyk, D., Pińskwar, I., & Choryński, A. (2022). Heat-related mortality in two regions of Poland: Focus on urban and rural areas during the most severe and long-lasting heatwaves. Atmosphere, 13(3), 390.

28. Graffy, P. M., Barrett, B. W., Horton, D. E., Kho, A. N., & Allen, N. B. (2025). Critical Temperature Thresholds for Identifying Vulnerability to Heat-Related Excess Cardiovascular Morbidity and Mortality. Journal of the American Heart Association, e046117.

29. Green, R. S., Basu, R., Malig, B., Broadwin, R., Kim, J. J., & Ostro, B. (2010). The effect of temperature on hospital admissions in nine California counties. International journal of public health, 55(2), 113–121.

30. Greenberg, J. H., Bromberg, J. I. L. L., Reed, C. M., Gustafson, T. L., & Beauchamp, R. A. (1983). The epidemiology of heat-related deaths, Texas--1950, 1970-79, and 1980. American Journal of Public Health, 73(7), 805–807.

31. Grömping, U. (2009). Variable importance assessment in regression: linear regression versus random forest. The American Statistician, 63(4), 308–319.

32. Guo, W., & Zhou, Z. Z. (2022). A comparative study of combining tree-based feature selection methods and classifiers in personal loan default prediction. Journal of Forecasting, 41(6), 1248–1313.

33. Harris, J. K., Hinyard, L., Beatty, K., Hawkins, J. B., Nsoesie, E. O., Mansour, R., & Brownstein, J. S. (2018). Evaluating the implementation of a twitter-based foodborne illness reporting tool in the city of St. Louis department of health. International journal of environmental research and public health, 15(5), 833.

34. Hastie, T., Tibshirani, R., & Friedman, J. (2009). The elements of statistical learning.

35. Healy, K. (2005). Heat wave: a social autopsy of disaster in Chicago. Imprints, 8(3), 283–289.

36. Hinkel, J. (2011). “Indicators of vulnerability and adaptive capacity”: towards a clarification of the science–policy interface. Global environmental change, 21(1), 198–208.

37. IPCC, 2023: Climate Change 2023: Synthesis Report. Contribution of Working Groups I, II and III to the Sixth Assessment Report of the Intergovernmental Panel on Climate Change [Core Writing Team, H. Lee and J. Romero (eds.)]. IPCC, Geneva, Switzerland, pp. 35–115, doi: 10.59327/IPCC/AR6-9789291691647.

38. Jeong, S., Lim, Y., Kang, Y., & Yi, C. (2024). Elucidating Uncertainty in Heat Vulnerability Mapping: Perspectives on Impact Variables and Modeling Approaches. International Journal of Environmental Research and Public Health, 21(7), 815.

39. Jiang, Z., Che, J., He, M., & Yuan, F. (2023). A CGRU multi-step wind speed forecasting model based on multi-label specific XGBoost feature selection and secondary decomposition. Renewable Energy, 203, 802–827.

40. Kaiser, R., Le Tertre, A., Schwartz, J., Gotway, C. A., Daley, W. R., & Rubin, C. H. (2007). The effect of the 1995 heat wave in Chicago on all-cause and cause-specific mortality. American journal of public health, 97(Supplement_1), S158-S162.

41. Kang, C., Park, C., Lee, W., Pehlivan, N., Choi, M., Jang, J., & Kim, H. (2020). Heatwave-related mortality risk and the risk-based definition of heat wave in South Korea: a nationwide time-series study for 2011–2017. International journal of environmental research and public health, 17(16), 5720.

42. Karpinska, L., Śmiech, S., Gouveia, J. P., & Palma, P. (2021). Mapping regional vulnerability to energy poverty in Poland. Sustainability, 13(19), 10694

43. Kephart, J. L., Sánchez, B. N., Moore, J., Schinasi, L. H., Bakhtsiyarava, M., Ju, Y.,…& Rodríguez, D. A. (2022). City-level impact of extreme temperatures and mortality in Latin America. Nature medicine, 28(8), 1700–1705.

44. Klinenberg, E. (1999). Denaturalizing disaster: A social autopsy of the 1995 Chicago heat wave. Theory and society, 28(2), 239–295.

45. Klinenberg, E. (2022). Heat wave: A social autopsy of disaster in Chicago. University of Chicago press.

46. Kollanus, V., Tiittanen, P., & Lanki, T. (2021). Mortality risk related to heatwaves in Finland–Factors affecting vulnerability. Environmental research, 201, 111503.

47. Li, F., Yigitcanlar, T., Nepal, M., Nguyen, K., Dur, F., & Li, W. (2025). Mapping heat vulnerability in Australian capital cities: A machine learning and multi-source data analysis. Sustainable Cities and Society, 119, 106079.

48. Lim, Y. H., Bell, M. L., Kan, H., Honda, Y., Guo, Y. L. L., & Kim, H. (2015). Economic status and temperature-related mortality in Asia. International journal of biometeorology, 59(10), 1405–1412.

49. Lin, S., Luo, M., Walker, R. J., Liu, X., Hwang, S. A., & Chinery, R. (2009). Extreme high temperatures and hospital admissions for respiratory and cardiovascular diseases. Epidemiology, 20(5), 738–746.

50. Lounici, K. (2008). Sup-norm convergence rate and sign concentration property of Lasso and Dantzig estimators.

51. Lüthi, S., Fairless, C., & Fischer, E. M. et al. (2023). Rapid increase in the risk of heat-related mortality. Nature Communications, 14, 4894. 10.1038/s41467-023-40599-x

52. Madrigano, J., Mittleman, M. A., Baccarelli, A., Goldberg, R., Melly, S., Von Klot, S., & Schwartz, J. (2013). Temperature, myocardial infarction, and mortality: effect modification by individual-and area-level characteristics. Epidemiology, 24(3), 439–446.

53. Mallen, E., Stone, B., & Lanza, K. (2019). A methodological assessment of extreme heat mortality modeling and heat vulnerability mapping in Dallas, Texas. Urban Climate, 30, 100528.

54. Manson, S., Schroeder, J., Van Riper, D., Knowles, K., Kugler, T., Roberts, F., & Ruggles, S. (2024). IPUMS National Historical Geographic Information System: Version 19.0 [Dataset]. Minneapolis, MN: IPUMS.

55. Manware, M., Dubrow, R., Carrión, D., Ma, Y., & Chen, K. (2022). Residential and race/ethnicity disparities in heat vulnerability in the United States. GeoHealth, 6, e2022GH000695. 10.1029/2022GH000695

56. Medina-Ramón, M., Zanobetti, A., Cavanagh, D. P., & Schwartz, J. (2006). Extreme temperatures and mortality: assessing effect modification by personal characteristics and specific cause of death in a multi-city case-only analysis. Environmental health perspectives, 114(9), 1331.

57. Michelozzi, P., De Donato, F., Bisanti, L., Russo, A., Cadum, E., DeMaria, M.,…& Perucci, C. A. (2005). The impact of the summer 2003 heat waves on mortality in four Italian cities. Euro surveillance: bulletin Europeen sur les maladies transmissibles= European communicable disease bulletin, 10(7), 11–12.

58. Muthukrishnan, R., & Rohini, R. L. A. S. S. O. (2016, October). LASSO: A feature selection technique in predictive modeling for machine learning. In 2016 IEEE international conference on advances in computer applications (ICACA) (pp. 18–20). Ieee.

59. National Weather Service, NOAA. (2022). The Heat Index Equation. Weather Prediction Center. Retrieved May 24, 2025, from https://www.wpc.ncep.noaa.gov/html/heatindex_equation.shtml

60. National Weather Service, NOAA. (n.d.). Vapor pressure calculator. National Oceanic and Atmospheric Administration. https://www.weather.gov/media/epz/wxcalc/vaporPressure.pdf

61. Naughton, M. P., Henderson, A., Mirabelli, M. C., Kaiser, R., Wilhelm, J. L., Kieszak, S. M.,…& McGeehin, M. A. (2002). Heat-related mortality during a 1999 heat wave in Chicago. American journal of preventive medicine, 22(4), 221–227.

62. Nguyen, T. T., Huang, J. Z., & Nguyen, T. T. (2015). Unbiased Feature Selection in Learning Random Forests for High-Dimensional Data. The Scientific World Journal, 2015(1), 471371.

63. Niu, Y., Li, Z., Gao, Y., & others. (2021). A systematic review of the development and validation of the heat vulnerability index: Major factors, methods, and spatial units. Current Climate Change Reports, 7(2), 87–97. 10.1007/s40641-021-00173-3

64. Ostro, B., Rauch, S., Green, R., Malig, B., & Basu, R. (2010). The effects of temperature and use of air conditioning on hospitalizations. American Journal of Epidemiology, 172(9), 1053–1061.

65. O’Neill, M. S., Zanobetti, A., & Schwartz, J. (2003). Modifiers of the temperature and mortality association in seven US cities. American journal of epidemiology, 157(12), 1074–1082.

66. O’Neill, M. S., Zanobetti, A., & Schwartz, J. (2005). Disparities by race in heat-related mortality in four US cities: the role of air conditioning prevalence. Journal of urban health, 82, 191–197.

67. Pillai, S. K., Noe, R. S., Murphy, M. W., Vaidyanathan, A., Young, R., Kieszak, S.,…& Wolkin, A. F. (2014). Heat illness: predictors of hospital admissions among emergency department visits—Georgia, 2002–2008. Journal of community health, 39(1), 90–98.

68. Potash, E., Brew, J., Loewi, A., Majumdar, S., Reece, A., Walsh, J.,…& Ghani, R. (2015). Predictive modeling for public health: Preventing childhood lead poisoning. Proceedings of the 21st ACM SIGKDD International Conference on Knowledge Discovery and Data Mining, 2039–2047.

69. Potash, E., Ghani, R., Walsh, J., Jorgensen, E., Lohff, C., Prachand, N., & Mansour, R. (2020). Validation of a machine learning model to predict childhood lead poisoning. JAMA network open, 3(9), e2012734.

70. Reid, C. E., O’neill, M. S., Gronlund, C. J., Brines, S. J., Brown, D. G., Diez-Roux, A. V., & Schwartz, J. (2009). Mapping community determinants of heat vulnerability. Environmental health perspectives, 117(11), 1730–1736.

71. Sadilek, A., Caty, S., DiPrete, L., Mansour, R., Schenk Jr, T., Bergtholdt, M.,…& Gabrilovich, E. (2018). Machine-learned epidemiology: real-time detection of foodborne illness at scale. NPJ digital medicine, 1(1), 36.

72. Sarofim, M. C., Saha, S., Hawkins, M. D., Mills, D. M., Hess, J., Horton, R., Kinney, P., Schwartz, J., & St. Juliana, A. (2016). Chapter 2: Temperature-related death and illness. In USGCRP (U.S. Global Change Research Program), The impacts of climate change on human health in the United States: A scientific assessment (pp. 43–69). 10.7930/J0MG7MDX

73. Schwartz, J. (2005). Who is sensitive to extremes of temperature?: A case-only analysis. Epidemiology, 16(1), 67–72.

74. Semenza, J. C., McCullough, J. E., Flanders, W. D., McGeehin, M. A., & Lumpkin, J. R. (1999). Excess hospital admissions during the July 1995 heat wave in Chicago. American journal of preventive medicine, 16(4), 269–277.

75. Semenza, J. C., Rubin, C. H., Falter, K. H., Selanikio, J. D., Flanders, W. D., Howe, H. L., & Wilhelm, J. L. (1996). Heat-related deaths during the July 1995 heat wave in Chicago. New England journal of medicine, 335(2), 84–90.

76. Shannon, E. M., Chen, L., Yuan, A., Chary, A., Gabrielian, S., Eisenman, D. P., & Washington, D. L. (2025). Extreme Heat, Social Factors, and Mortality Among California Veterans With Cardiometabolic Disease. JAMA Network Open, 8(11), e2545524–e2545524.

77. Shi, X., Wong, Y. D., Li, M. Z. F., Palanisamy, C., & Chai, C. (2019). A feature learning approach based on XGBoost for driving assessment and risk prediction. Accident Analysis & Prevention, 129, 170–179.

78. Spangler, K. R., Weinberger, K. R., & Wellenius, G. A. (2019). Suitability of gridded climate datasets for use in environmental epidemiology. Journal of exposure science & environmental epidemiology, 29(6), 777–789.

79. Speiser, J. L., Miller, M. E., Tooze, J., & Ip, E. (2019). A comparison of random forest variable selection methods for classification prediction modeling. Expert systems with applications, 134, 93–101.

80. Stafoggia, M., Forastiere, F., Agostini, D., Biggeri, A., Bisanti, L., Cadum, E.,…& Perucci, C. A. (2006). Vulnerability to heat-related mortality: a multicity, population-based, case-crossover analysis. Epidemiology, 17(3), 315–323.

81. Stang, A., Standl, F., Kowall, B., Brune, B., Böttcher, J., Brinkmann, M.,…& Jöckel, K. H. (2020). Excess mortality due to COVID-19 in Germany. Journal of Infection, 81(5), 797–801.

82. Sun, S., Wang, H., Cheng, C., Chang, Z., & Huang, D. (2017, December). PCA-based heart sound feature generation for a ventricular septal defect discrimination. In 2017 14th International Computer Conference on Wavelet Active Media Technology and Information Processing (ICCWAMTIP) (pp. 128-133). IEEE.

83. Tate, E. (2012). Social vulnerability indices: a comparative assessment using uncertainty and sensitivity analysis. Natural hazards, 63(2), 325–347.

84. Theng, D., & Bhoyar, K. K. (2024). Feature selection techniques for machine learning: a survey of more than two decades of research. Knowledge and Information Systems, 66(3), 1575–1637.

85. Thompson, R., Landeg, O., Kar-Purkayastha, I., Hajat, S., Kovats, S., & O’connell, E. (2022). Heatwave mortality in summer 2020 in England: an observational study. International journal of environmental research and public health, 19(10), 6123.

86. Thornton, M. M., Shrestha, R., Wei, Y., Thornton, P. E., & Kao, S.-C. (2022). Daymet: Daily surface weather data on a 1-km grid for North America, Version 4 R1 [Data set]. ORNL DAAC. 10.3334/ORNLDAAC/2129

87. Thornton, P. E., Running, S. W., & White, M. A. (1997). Generating surfaces of daily meteorological variables over large regions of complex terrain. Journal of hydrology, 190(3-4), 214–251.

88. Tibshirani, R. (1996). Regression shrinkage and selection via the lasso. Journal of the Royal Statistical Society: Series B (Methodological), 58, 267–288.

89. Toloo, G. S., Guo, Y., Turner, L., Qi, X., Aitken, P., & Tong, S. (2014). Socio-demographic vulnerability to heatwave impacts in Brisbane, Australia: a time series analysis. Australian and New Zealand journal of public health, 38(5), 430–435.

90. Uejio, C. K., Wilhelmi, O. V., Golden, J. S., Mills, D. M., Gulino, S. P., & Samenow, J. P. (2011). Intra-urban societal vulnerability to extreme heat: the role of heat exposure and the built environment, socioeconomics, and neighborhood stability. Health & place, 17(2), 498–507.

91. U.S. Global Change Research Program (USGCRP). (2023). Fifth National Climate Assessment (A. R. Crimmins, C. W. Avery, D. R. Easterling, K. E. Kunkel, B. C. Stewart, & T. K. Maycock, Eds.). U.S. Global Change Research Program. https://nca2023.globalchange.gov/

92. Walton, D., & Hall, A. (2018). An assessment of high-resolution gridded temperature datasets over California. Journal of Climate, 31(10), 3789–3810.

93. Wang, S., Cai, W., Tao, Y., Sun, Q. C., Wong, P. P. Y., Huang, X., & Liu, Y. (2023). Unpacking the inter-and intra-urban differences of the association between health and exposure to heat and air quality in Australia using global and local machine learning models. Science of The Total Environment, 871, 162005. 10.1016/j.scitotenv.2023.162005

94. Whitman, S., Good, G., Donoghue, E. R., Benbow, N., Shou, W., & Mou, S. (1997). Mortality in Chicago attributed to the July 1995 heat wave. American Journal of public health, 87(9), 1515–1518.

95. Wolf, T., McGregor, G., & Analitis, A. (2014). Performance assessment of a heat wave vulnerability index for greater London, United Kingdom. Weather, climate, and society, 6(1), 32–46.

96. World Health Organization. (2024, May 28). Climate change, heat and health. https://www.who.int/news-room/fact-sheets/detail/climate-change-heat-and-health

97. World Meteorological Organization. (2023, September 21). Climate change and heatwaves. https://wmo.int/content/climate-change-and-heatwaves

98. Xu, Z., Tong, S., Cheng, J., Crooks, J. L., Xiang, H., Li, X.,…& Hu, W. (2019). Heatwaves and diabetes in Brisbane, Australia: a population-based retrospective cohort study. International Journal of epidemiology, 48(4), 1091–1100.

99. Yin, P., Chen, R., Wang, L., Liu, C., Niu, Y., Wang, W.,…& Kan, H. (2018). The added effects of heatwaves on cause-specific mortality: a nationwide analysis in 272 Chinese cities. Environment international, 121, 898–905.

100. Zanobetti, A., O’Neill, M. S., Gronlund, C. J., & Schwartz, J. D. (2013). Susceptibility to mortality in weather extremes: effect modification by personal and small-area characteristics. Epidemiology, 24(6), 809–819.

101. Zhang, L., Zhang, Z., Ye, T., Zhou, M., Wang, C., Yin, P., & Hou, B. (2018). Mortality effects of heat waves vary by age and area: a multi-area study in China. Environmental Health, 17(1), 54.

102. Zhao, L., Qing, S., Bai, J., Hao, H., Li, H., Shi, Y.,…& Yang, R. (2023). A hybrid optimized model for predicting evapotranspiration in early and late rice based on a categorical regression tree combination of key influencing factors. Computers and Electronics in Agriculture, 211, 108031.

103. Zheng, H., Yuan, J., & Chen, L. (2017). Short-term load forecasting using EMD-LSTM neural networks with a Xgboost algorithm for feature importance evaluation. Energies, 10(8), 1168.

104. Zhou, H., Ma, L., Niu, X., Xiang, Y., Chen, J., Su, Y.,…& Wu, Q. (2024). A novel hybrid model combined with ensemble embedded feature selection method for estimating reference evapotranspiration in the North China Plain. Agricultural Water Management, 296, 108807.

