## Supplemental Material for "Integrating Machine Learning-Based Variable Selection into Heat Vulnerability Index Design"

**Supplemental Figure S1.** Interactive map of the Community Areas (CAs) in Chicago. The interactive map shows the spatial distribution and area size of the 77 CAs in Chicago. Hover the mouse to display the CA name and area (km²). The interactive function is implemented based on the OpenStreetMap base map.

We first project the original shapefile data onto the UTM zone 16N suitable for Chicago (EPSG:32616) to calculate the area in square kilometers, and then reproject the data back to EPSG:4326 for visualization. Data source: Chicago Data Portal (https://data.cityofchicago.org). Geospatial analysis based on GeoPandas and Folium libraries.

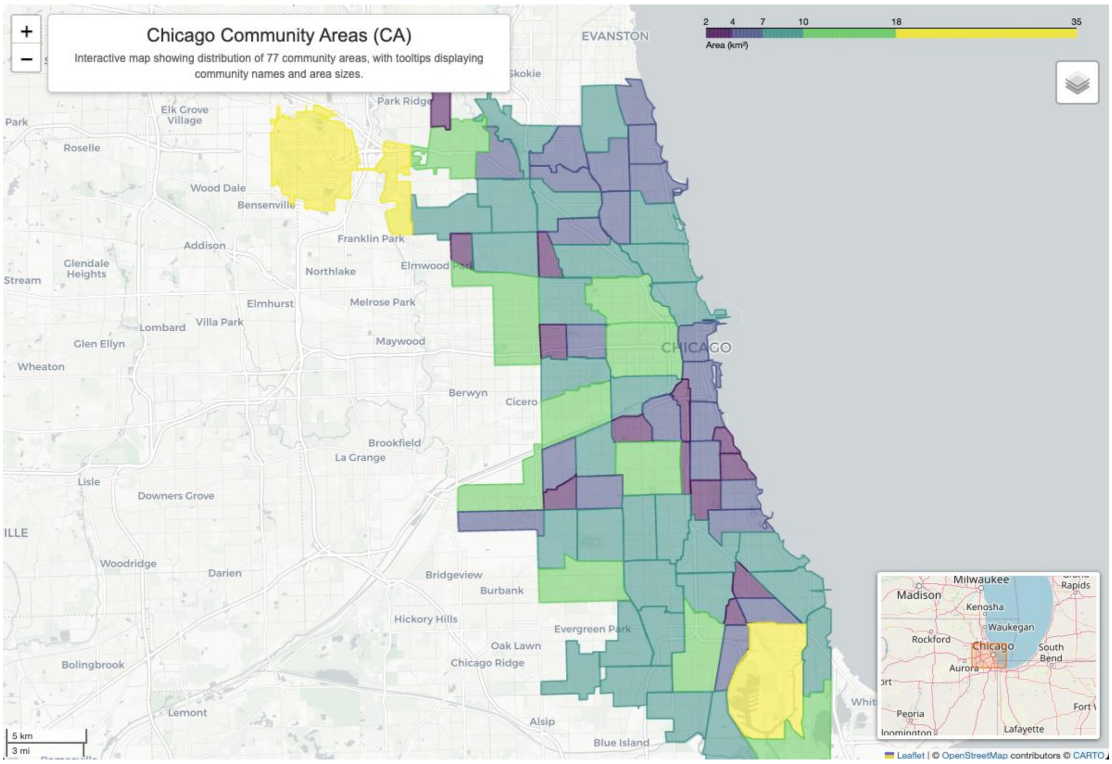


**Supplemental Table S1.** Crosswalk of heat vulnerability indicators in Reid et al. (2009) and this study.

| Category | Original indicator (Reid et al. 2009) | Indicator used in this study | Definition as used in this study |
|---|---|---|---|
| Diabetes prevalence | Diabetes | Diabetes Prevalence | Percentage of adults who reported having ever been diagnosed with diabetes by a doctor or other health professional, excluding pre-diabetes and gestational diabetes. |
| Demographic and socioeconomic variables | Race other than White | Black (Non-Hispanic) | Proportion of Non-Hispanic Black residents |
| | | Hispanic or Latino (of Any Race) | Proportion of Hispanic or Latino residents |
| | Age > 65 years | Age above 65 | Proportion of the population aged 65 and over |
| | Live alone | Living Alone | Proportion of individuals living alone |
| | Age > 65 living alone | Age above 65 and Living Alone | Proportion of older adults living alone |
| | Below poverty line | Poverty Rate | Proportion of residents living in households below the federal poverty threshold |
| | Less than high school diploma | Less than High School Diploma | Proportion of the population aged 25 years and over with less than a high school diploma |
| Land cover | Not Green Space | Not Green Space | Proportion of area not classified by green space |
| Air conditioning | No central AC<br>No AC of any kind | No AC Access | Percentage of households reporting no air conditioning equipment is used in the home. |

**Supplemental Figure S2.** Correlations among heat vulnerability indicators. The heatmap demonstrates the Pearson correlation coefficients for the heat vulnerability indicators considered in this study, all of which are below 0.75.

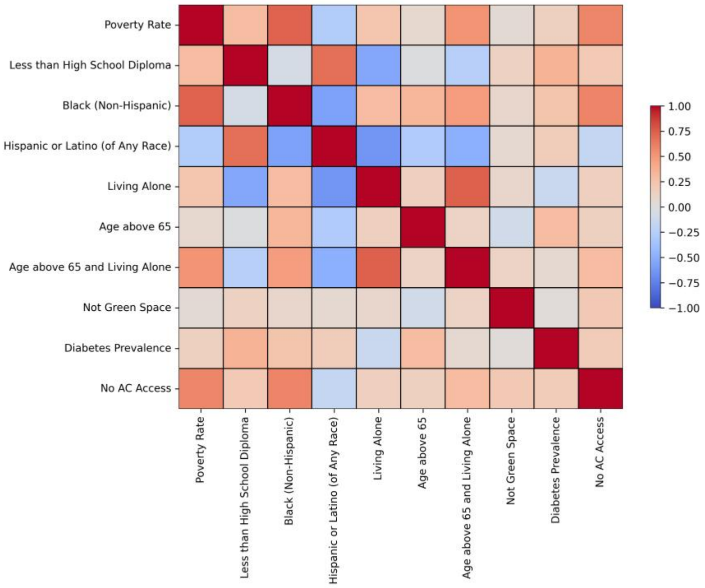


**Supplemental Figure S3.** Time series of heat-related excess mortality in Chicago, 1993–2019. (a) Annual heat-related excess mortality; (b) Average daily heat-related excess mortality. Linear trend lines are shown to illustrate long-term temporal patterns, with slope estimates and corresponding p-values.

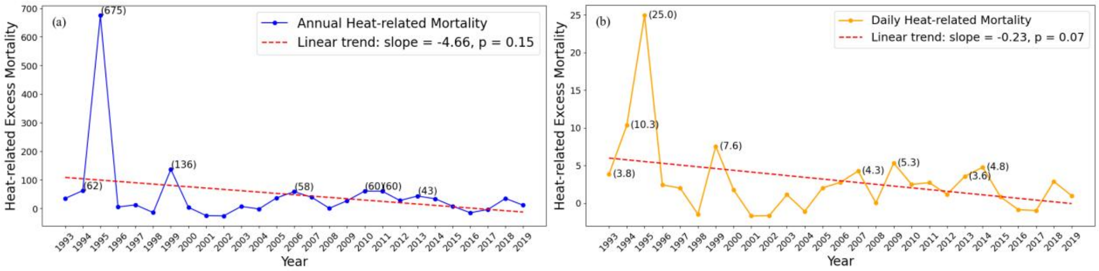


**Supplemental Figure S4.** Time series of heat-related excess mortality in Chicago, 1993–2019 (excluding 1995). (a) Annual heat-related excess mortality; (b) Average daily heat-related excess mortality. Mean and median reference lines are shown, and values exceeding the mean are annotated.

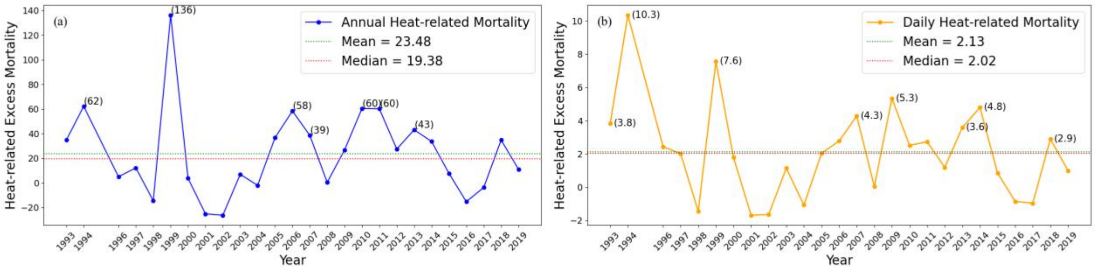

**Supplemental Figure S5.** Time series of heat-related excess mortality in Chicago, 1993–2019 (excluding 1995). (a) Annual heat-related excess mortality; (b) Average daily heat-related excess mortality. Linear trend lines are shown to illustrate long-term temporal patterns, with slope estimates and corresponding p-values.

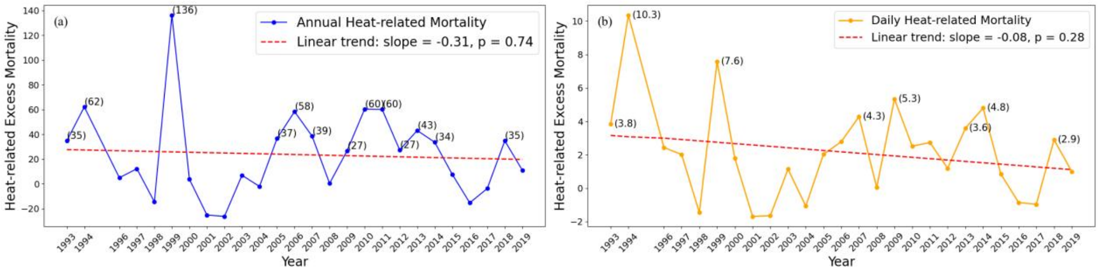


**Supplemental Table S2.** Simple linear regression result. Significance threshold: p < 0.05. Ranked by the absolute values of beta coefficients.

| Indicator | Coefficient | p-value |
|---|---|---|
| Poverty Rate | 0.46 | <0.005 |
| No AC Access | 0.38 | <0.005 |
| Black (Non-Hispanic) | 0.33 | <0.005 |
| Age above 65 and Living Alone | 0.30 | 0.01 |
| Age above 65 | 0.23 | 0.04 |
| Less than High School Diploma | 0.18 | 0.13 |
| Hispanic or Latino (of Any Race) | -0.16 | 0.16 |
| Living Alone | 0.15 | 0.21 |
| Not Green Space | -0.05 | 0.65 |
| Diabetes Prevalence | 0.04 | 0.76 |

**Supplemental Table S3.** Polynomial regression result. Indicators were selected based on the joint F-test of polynomial terms with a significance threshold of p-value < 0.05. The indicators are ranked by the F-statistics.

| Indicator | Joint F-test (degree = 2) | | Joint F-test (degree = 3) | |
|---|---|---|---|---|
| | F-statistic | p-value | F-statistic | p-value |
| Poverty Rate | 10.44 | <0.005 | 7.97 | <0.005 |
| No AC Access | 9.94 | <0.005 | 6.74 | <0.005 |
| Black (Non-Hispanic) | 4.90 | 0.010 | 3.73 | 0.015 |
| Age above 65 and Living Alone | 4.25 | 0.018 | 2.86 | 0.043 |
| Age above 65 | 4.18 | 0.019 | 2.85 | 0.043 |
| Less than High School Diploma | 2.76 | 0.070 | 2.75 | 0.049 |
| Not Green Space | 1.06 | 0.351 | 0.86 | 0.466 |
| Hispanic or Latino (of Any Race) | 0.95 | 0.391 | 0.70 | 0.552 |
| Living Alone | 0.93 | 0.398 | 0.63 | 0.601 |
| Diabetes Prevalence | 0.70 | 0.499 | 0.62 | 0.602 |

**Supplemental Table S4.** Lasso Regression result. Indicators with non-zero coefficients are selected and ranked by the absolute values of their coefficients.

| Indicator | Coefficient |
|---|---|
| Poverty Rate | 0.095 |
| Age above 65 | 0.028 |
| Not Green Space | -0.023 |
| Diabetes Prevalence | -0.009 |
| Less than High School Diploma | 0.005 |
| No AC Access | 0 |
| Black (Non-Hispanic) | 0 |
| Age above 65 and Living Alone | 0 |
| Hispanic or Latino (of Any Race) | 0 |
| Living Alone | 0 |

**Supplemental Table S5.** Random Forest result. The importance reflects the indicator's contribution to predicting heat-related excess mortality. The standard deviation of the importance reflects the variability of its estimate across different trees, with a smaller standard deviation indicating more stable and reliable importance of the indicator. Indicators are ranked by their importance, and those with importance scores greater than 0.01 are retained.

| Indicator | Importance | Standard Deviation |
|---|---|---|
| Poverty Rate | 0.025 | 0.031 |
| No AC Access | 0.018 | 0.026 |
| Less than High School Diploma | 0.014 | 0.021 |
| Age above 65 | 0.012 | 0.018 |
| Age above 65 and Living Alone | 0.011 | 0.022 |
| Hispanic or Latino (of Any Race) | 0.010 | 0.018 |
| Black (Non-Hispanic) | 0.008 | 0.016 |
| Living Alone | 0.007 | 0.015 |
| Diabetes Prevalence | 0.004 | 0.009 |
| Not Green Space | 0.003 | 0.007 |

**Supplemental Figure S6.** XGBoost result. Indicators are ranked by their feature importance (gain), and those contributing to at least 80% of the cumulative importance were retained.

| Indicator | Feature Importance | Value | Cumulative Importance |
|---|---|---|---|
| No AC Access | | 0.300 | 0.300 |
| Diabetes Prevalence | | 0.177 | 0.477 |
| Not Green Space | | 0.116 | 0.593 |
| Poverty Rate | | 0.106 | 0.699 |
| Black (Non-Hispanic) | | 0.099 | 0.798 |
| Age above 65 | | 0.082 | 0.880 |
| Less than High School Diploma | | 0.073 | 0.953 |
| Hispanic or Latino (of Any Race) | | 0.047 | 1.000 |
| Living Alone | | 0.000 | 1.000 |
| Age above 65 and Living Alone | | 0.000 | 1.000 |

**Supplemental Figure S7.** Spatial distribution of heat-related excess mortality rate across communities in Chicago, 1993–2019. Heat days are defined as at least two consecutive days with the daily maximum Heat Index exceeding 110°F (43.3°C). (a) Map of the average annual heat-related excess mortality rate (range: -1.3 to 6.5 per 100,000; mean: 1.47 per 100,000). Spatial autocorrelation is statistically significant (Moran's I = 0.351, p = 0.001). (b) Map of the average daily heat-related excess mortality (range: -0.48 to 2.16 per 100,000; mean: 0.50 per 100,000). Spatial autocorrelation is statistically significant (Moran's I = 0.343, p = 0.001).

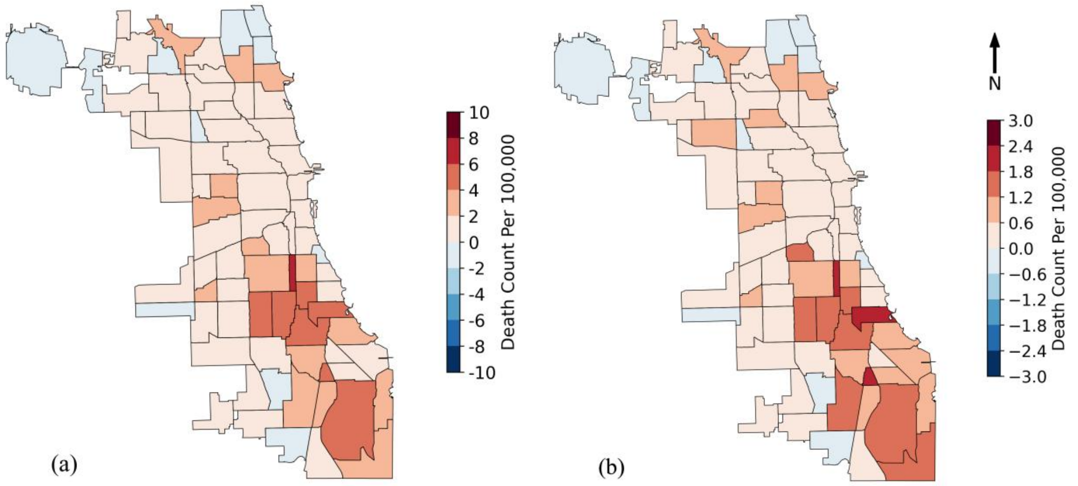
